# Exome wide association study for blood lipids in 1,158,017 individuals from diverse populations

**DOI:** 10.1101/2024.09.17.24313718

**Authors:** Satoshi Koyama, Zhi Yu, Seung Hoan Choi, Sean J. Jurgens, Margaret Sunitha Selvaraj, Derek Klarin, Jennifer E. Huffman, Shoa L. Clarke, Michael N. Trinh, Akshaya Ravi, Jacqueline S. Dron, Catherine Spinks, Ida Surakka, Aarushi Bhatnagar, Kim Lannery, Whitney Hornsby, Scott M. Damrauer, Kyong-Mi Chang, Julie A Lynch, Themistocles L. Assimes, Philip S. Tsao, Daniel J. Rader, Kelly Cho, Gina M. Peloso, Patrick T. Ellinor, Yan V. Sun, Peter WF. Wilson, Million Veteran Program, Pradeep Natarajan

## Abstract

Rare coding alleles play crucial roles in the molecular diagnosis of genetic diseases. However, the systemic identification of these alleles has been challenging due to their scarcity in the general population. Here, we discovered and characterized rare coding alleles contributing to genetic dyslipidemia, a principal risk for coronary artery disease, among over a million individuals combining three large contemporary genetic datasets (the Million Veteran Program, n = 634,535, UK Biobank, n = 431,178, and the All of Us Research Program, n = 92,304) totaling 1,158,017 multi-ancestral individuals. Unlike previous rare variant studies in lipids, this study included 238,243 individuals (20.6%) from non-European-like populations.

Testing 2,997,401 rare coding variants from diverse backgrounds, we identified 800 exome-wide significant associations across 209 genes including 176 predicted loss of function and 624 missense variants. Among these exome-wide associations, 130 associations were driven by non-European-like populations. Associated alleles are highly enriched in functional variant classes, showed significant additive and recessive associations, exhibited similar effects across populations, and resolved pathogenicity for variants enriched in African or South-Asian populations. Furthermore, we identified 5 lipid-related genes associated with coronary artery disease *(RORC, CFAP65, GTF2E2, PLCB3, and ZNF117)*. Among them, *RORC* is a potentially novel therapeutic target through the down regulation of LDLC by its silencing.

This study provides resources and insights for understanding causal mechanisms, quantifying the expressivity of rare coding alleles, and identifying novel drug targets across diverse populations.

---

Family-based discovery and characterization of rare coding alleles causative of familial hypercholesterolemia (FH) have yielded important insights for coronary artery disease (CAD), the leading cause of premature mortality among adults^1,2^. While FH is associated with a heightened risk for early-onset CAD, early intervention using lipid-lowering medications can considerably mitigate this risk, suppressing cumulative exposure to continuously high levels of low-density lipoprotein cholesterol (LDLC)^3,4^. However, FH remains substantially underdiagnosed and undertreated^4–7^. This highlights the need for increased efforts to identify and characterize pathogenic variants associated with FH.

Additionally, like other Mendelian conditions, population-based genetic analyses have often shown that expressivity (continuous effects on lipid levels) and penetrance (likelihood of CAD) may not be sufficiently high for some previously implicated pathogenic variants relative to initial descriptions in family-based studies^8–13^. As rare Mendelian alleles are increasingly returned to asymptomatic individuals through screening or secondary reporting^14–16^, allele-specific prognosis is increasingly important.

Furthermore, clinically curated variants are enriched among individuals genetically similar to European reference populations, reflecting biases in accumulated knowledge and data. In contrast, variants associated with non-European reference populations are more likely to be reclassified^17^, susceptible to population-related biased filters,^18^ underdiagnosed due to limited data availability^19^.

To address these challenges, we assembled a large-scale, finely imputed/sequenced dataset encompassing lipid measures from over a million individuals combining the Million Veteran Program (MVP)^20^, UK Biobank (UKB)^21^, and the All of Us Research Program (AOU) cohorts, which included more than 230,000 that are genetically similar to non-European reference populations - historically underrepresented in genomic research. This diverse dataset allowed us to identify and characterize rare coding variant associations with blood lipids and validate the generalizability across populations. The summary of the estimated effects will provide a resource for further functional assessment and clinical utility.

## Study population

We generated a large-scale clinical genetic dataset by imputing MVP (634,535 individuals) to TOPMed imputation reference panel version r2^22^, which includes 308,107,085 variants from 97,256 individuals representing diverse populations. Combined with whole exome sequence (WES) data in UKB (431,178 individuals)^23^ and whole genome sequence (WGS) data in AOU (92,304 individuals)^24^, we generated a cohort of 1,158,017 individuals, including 238,243 (20.57%) from non-European populations (Fig. 1a, Supplementary Table 1). Large scale imputation reference panel including diverse populations allowed us to impute rare variants with high accuracy comparable to sequenced data (Extended Data Figures 1a-1c, Supplementary Notes I).

**Fig. 1.**
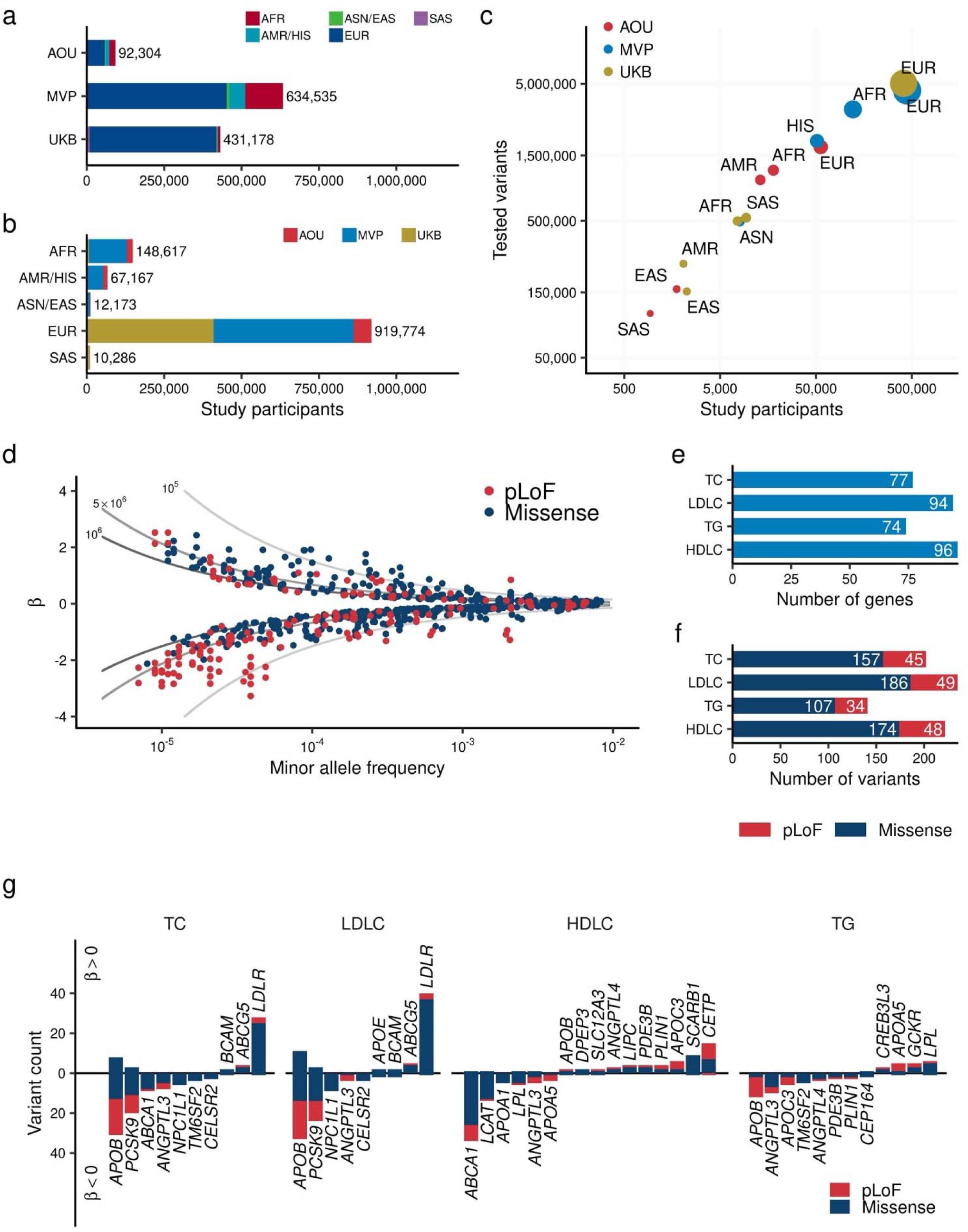
Exome wide association study for blood lipids over one million individuals. **a.** and **b.** Overview of the study. The number of individuals included in the analysis by study (**a**) and by population (**b**). **c.** Correlation between the number of individuals and identified variants in the target region. The horizontal axis shows the number of individuals in each population by study. The vertical axis shows the number of variants identified in the corresponding population. The size of point is proportional to the number of individuals. **d.** Distribution of effect sizes for exome-wide significant associations is shown. Each dot represents a variant-trait pair with significant association in this study (Methods). All four blood lipids are plotted. The horizontal axis indicates the minor allele frequency, while the vertical axis displays the effect size for each allele from the regression model (β), with the unit of effect size normalized to the standard deviations of blood lipids. The lines represent the statistical power of 80% at sample sizes of one million (dark gray), 500,000 (medium gray), and 100,000 (light gray) individuals. **c.** Minor allele frequency of associated variants by variant impact. The rectangles illustrate the interquartile range of the minor allele frequencies, with the bottom and top edges representing the first and third quartiles, respectively. The line inside the rectangle denotes the median and the whiskers extend from the quartiles to the smallest and largest observed values, within a distance no greater than 1.5 times the interquartile range. **d.** Direction of the effects for associated variants. Variants positively associated with the blood lipids are displayed on the positive side of the vertical axis. The height of each bar represents the number of variants in that category. Bar colors indicate variant classes, with blue for missense variants and red for pLoF variants. AFR, African-like population; AMR, Admixed-American-like population; ASN, Asian-like population; EAS, East-Asian-like population; EUR, European-like population; HIS, Hispanic-like population; SAS, South-Asian-like population; TC, Total Cholesterol; LDLC, Low Density Lipoprotein Cholesterol; HDLC, High Density Lipoprotein Cholesterol; TG, Triglycerides; pLoF, predicted Loss of Function; EWS, Exome Wide Significance.

## Variant identification

We curated the variants with minor allele count (MAC) ≥ 5 detected in ± 50 base pairs of exome target region used in UKB-WES (Methods). Annotation using 19,603 protein coding transcripts identified 214,000 predicted Loss of Function (pLoF, stop gain, frameshift insertion/deletion, and canonical splice site), and 2,766,489 missense variants [missense single nucleotide variant (SNV), and in-frame insertion/deletion, Supplementary Table 2]. These variants covered 1.72% of all possible pLoF SNVs and 3.72% of all possible missense SNVs (Extended Data Figure 1d, Supplementary Table 3, Supplementary Notes II).

In addition, using the Splice-AI algorithm^25^, we detected 23,523 putatively cryptic splice variants [variants associated with donor/acceptor-gain, donor/acceptor-loss in distant position from canonical splice site with Delta Score > 0.8. Supplementary Notes III]. We re-classified these cryptic splice variants as pLoF and included them in association analyses. In total we identified 237,523 pLoF variants and 2,759,878, missense variants in this study (Supplementary Table 4). The MVP and AOU study populations, including diverse populations, effectively increases the variety of variants included in this study (Extended Data Figures 1b and 1c).

We identified at least one testable pLoF in 89.20% (17,486/19,603) of assessed transcripts and a missense variant in 95.30% (18,682/19,603). Among them, 71.29% (12,465/17,486) and 99.37% (18,666/18,682) of transcript had at least one pLoF or missense variant with 80% statistical power to detect effect size of one standard deviation (SD) of phenotypes per allele (Extended Data Figures 1e and 1f, Supplementary Table 5, Supplementary Notes IV).

## Association analysis

We tested linear associations of the imputed/sequenced genotypes of rare (5 ≤ MAC and MAF_POPMAX_ < 1%) pLoF variants or missense variants with blood lipids [total cholesterol (TC), LDLC, high-density lipoprotein cholesterol (HDLC) triglycerides (TG)] using an additive model stratified by population groups (4 from MVP, 5 from UKB, and 5 from AOU, Fig. 1a, Supplementary Table 1, Methods) followed by fixed effects meta-analysis including 14 population groups. In total, we tested 11,226,703 variant-phenotype combinations in the additive model. The highest Lambda GC in four tested traits was 1.025 for HDLC indicating suitable calibration (Extended Data Figure 2a). In addition, we conducted recessive model analysis for 233,971 variant-phenotype combinations with 5 ≤ minor homozygote counts and minor homozygote frequency < 1%. Exome-wide significance (EWS) was defined as *P* < 4.4 × 10^−9^ [0.05/(11,226,703 + 233,971)].

We identified 800 additive EWS associations in 184 loci (202 associations in 45 loci for TC; 235 in 48 for LDLC; 222 in 47 for HDLC; and 141 in 44 for TG, Extended Data Figure 2b, Supplementary Table 6), and 110 recessive EWS associations (Supplementary Table 7). The additive signals included 176 pLoF associations across 40 genes and 623 missense associations across 193 genes (Figs. 1b and 1c) often with multiple associations per gene (Fig. 1d).

We observed significant enrichment of EWS variants in pLoF or missense variants compared to synonymous/non-coding variants [odds ratio (OR)_EWS/Non-EWS_ = 6.33, 95% Confidence Interval (CI) = 5.02 – 7.91, *P =* 6.0 × 10^−41^ for pLoF variants, and 2.27 (1.98 – 2.60), *P =* 8.9 × 10^−32^ for missense variants]. One of the strongest signals was the *APOB* pLoF variant (p.M3438X), which altered LDLC by −3.14 SD per allele (or −103.53 mg/dL per allele, mean LDLC was 57.9mg/dL for 5 carriers, and 145mg/dL for 409,041 non-carriers, Extended Data Figure 2c).

To assess the replicability of the results, we compared the effect sizes with a previous independent microarray-based rare-variant study for blood lipids (Lu et al., *Nat Genet* 2017, N = 358,251)^26^. In the replication dataset, we identified 48.4% (387/800) of EWS associations. 99.7% (386/387) of these variants showed directional concordance and 41.9% (162/387) showed significant association in the replication dataset (*P* < 0.05/387). For the eleven variants found in ten novel loci identified in this study, we found 81.8% (9/11) associations in the replication dataset. All 9 of these associations showed concordant effect direction and 5 of these showed nominal association (*P* < 0.05, Extended Data Figure 2d) in the replication dataset.

## Variant function predicts phenotype expressivity

To gain further insights into genetic associations and variant functions, we employed existing *in silico* methods for predicting variant functionality. For pLoF variants, we utilized the LOFTEE^27^ plugin in VEP^28^ and identified 163,643 ‘high-confidence’ pLoF variants (87.7% of pLoF variants). For missense variants, we applied 29 *in silico* deleterious prediction algorithms^29^, from which we derived an ensembled Missense Score (MiS, Methods) and grouped them into bins ([0, 0.5], (0.5, 0.7], (0.7, 0.9], and (0.9, 1], where deleteriousness increases with increasing value. Supplementary Tables 8 and 9). We observed strong linear relationships across variant deleteriousness, lower allele frequencies, and phenotype association (Fig. 2a, Supplementary Table 10). Notably, high-confidence pLoF, deleterious missense variants with a MiS (0.9, 1.0], and (0.7, 0.9] exhibited similarly constrained low MAF (median MAF 0.0023%, 0.0021%, and 0.0024%, respectively) and were more likely to be EWS [OR_EWS/Non-EWS_ = 7.24 (95% CI 5.66 – 91.8) and *P* = 5.2 × 10^−40^ for pLoF; OR = 11.61 (7.02 – 18.15) and *P* = 6.2 × 10^−15^ for MiS (0.9, 1.0]; OR = 5.02 (3.87 – 6.43) and *P* = 1.9 × 10^−26^ for MiS (0.7, 0.9]]. Furthermore, the cryptic splice variants exhibited a similar level of constraint (median MAF 0.0028%) and were equally enriched for EWS [OR_EWS/Non-EWS_ = 5.96 (95% CI 2.71 – 11.42), *P* = 3.1 × 10^−5^]. As an example, among the 53 pLoF variants observed in *APOB*, 19 were EWS, but these associated variants were depleted in the last exon (Fig. 2b) and predicted as “low-confident” pLoF.

**Fig. 2.**
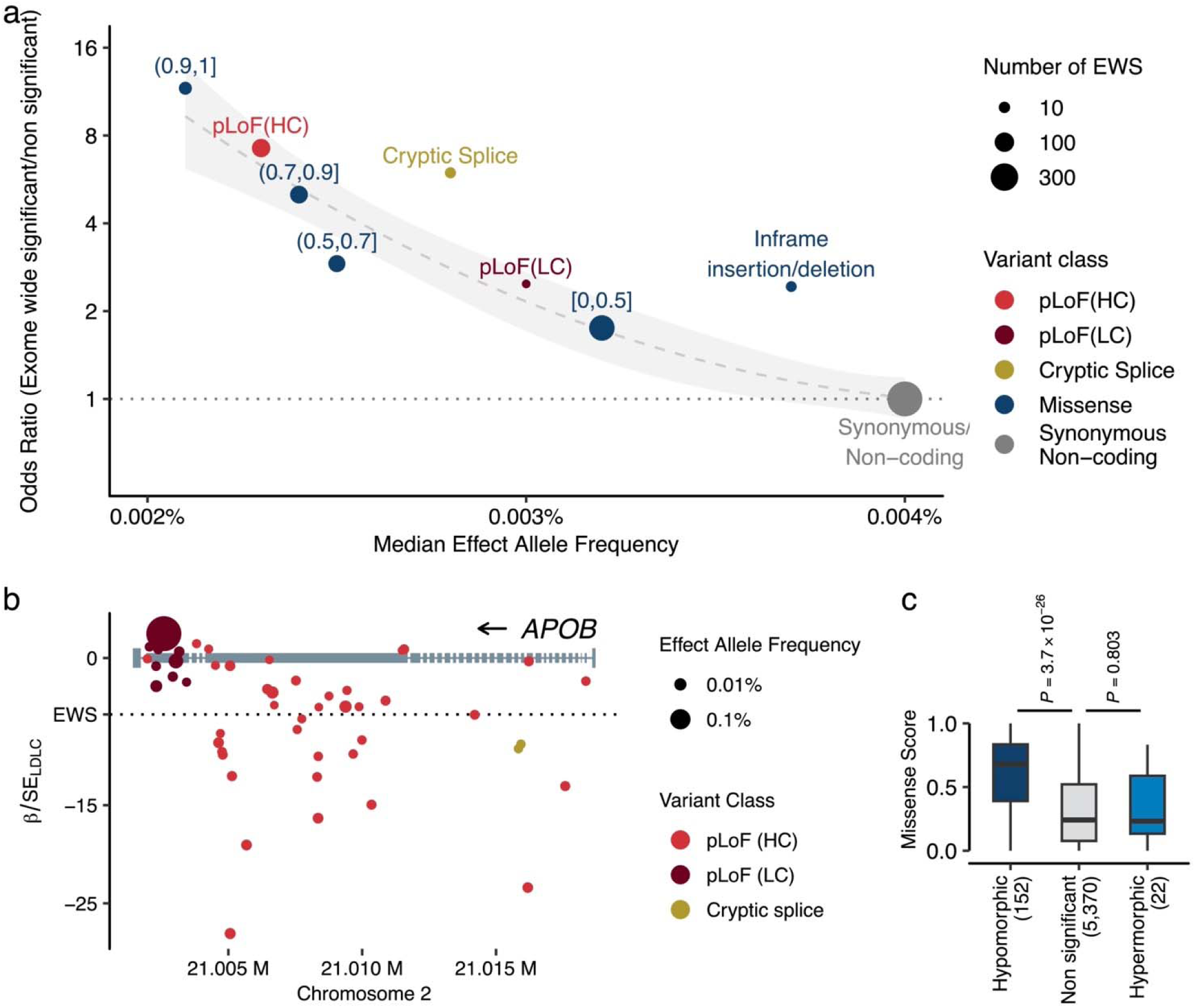
Different expressivity of rare coding variants by variant classes. **a.** Variant deleteriousness, constraints, and statistical associations. The panel represents variant classes as pLoF (red), Missense (blue), and Synonymous/Non-coding (gray, used as reference). The ranges associated with the blue points depict the Missense Score for missense variants. We computed the Missense Score for missense single nucleotide variants by using 29 in-silico deleteriousness prediction algorithms. The score was calculated as the number of deleterious predictions divided by the number of available algorithms for each variant, with values ranging from 0 to 1 (Methods). Based on the Missense Scores, missense variants were grouped into bins. pLoF variants were grouped by LOFTEE predictions. The horizontal axis indicates the median minor allele frequency for each variant class, while the vertical axis shows the odds ratios of EWS to non-EWS variants in reference to Synonymous/Non-coding variants. Odds ratios were estimated by Fisher’s Exact test. Circle size corresponds to the number of variants achieving EWS in each variant class. The dashed curve is the estimated line, and the shaded area is its 95% confidence interval. **b.** Penetrance of pLoF variants in the *APOB*. Gray rectangles represent the *APOB* gene model. Circles correspond to genetic variants examined in this study, with circle size denoting effect allele frequency, and color signifying variant class. The horizontal axis outlines genomic coordinates (hg38), whereas the vertical axis indicates Z-values (Beta/Standard Error) for LDLC association calculated by liner mixed model (Methods). **c.** Different distributions of Missense Scores (See above) observed in hypermorphic and hypomorphic variants. The box plot displays the distribution of Missense Scores for Missense variants within genes that have at least one EWS association by pLoF. A hypomorphic variant is defined as having the same directional association with EWS pLoF association. The *P*-values were calculated by two-sided Wilcoxon’s rank-sum test. The *P*-values were not adjusted for multiple testing correction. Conversely, a hypermorphic variant is defined as having an opposite directional association to EWS. pLoF, predicted Loss of Function; HC, High Confidence; LC, Low Confidence; EWS, Exome Wide Significance; LDLC, Low Density Lipoprotein Cholesterol.

## Distinguishing hypomorphic and hypermorphic missense variants

Multiple EWS pLoF associations allowed us to assess the effect directions of gene silencing in 23 gene-phenotype pairs (Fig. 1d). These included 128 pLoF variant-phenotype pairs, and all exhibited consistent effect directions except for a cryptic splice variant in *CETP* and HDLC. 87% (239/275) of missense variants showed concordant effect directions with pLoF variants in the same genes (hypomorphic variants). However, 36 associations in 10 genes were found to have opposite effect direction to pLoF variants (hypermorphic variants, Supplementary Table 6). Some previously discovered hypermorphic variants included *PCSK9* [p.R469W^30^, p.R496W^31^] and *APOB* [p.R3527Q] but most are newly discovered hypermorphic variants. One such example is *LDLR* p.S849L which showed strong negative association with LDLC [β = −1.07 (SE 0.087), *P* = 3.6×10^−34^] indicating gain-of-function. Another example is *APOB* p.G4395S which is of higher MAF in the African-like population [β_MVP-AFR_ = 0.433 (SE 0.080), β_UKB-AFR_ = 0.775 (0.253)]. While MiS was an important factor in predicting hypomorphic associations, it did not predict hypermorphic associations (Fig. 2c).

## Cryptic splicing variants as novel candidates for loss-of-function

For all identified variants in this study, we predicted the variant’s potential for splice site disruption/creation using Splice-AI^25^ and derived a Delta Score (DS) – a numeric score ranging 0 – 1 (Supplementary Notes III). The score distribution was sparse and only 0.598% (58,402/9,399,797) of variants had high DS (> 0.8). 43.5% of variants with high DS were not located in the canonical splice sites (Extended Data Figure 3a). We observed a strong enrichment of cryptic splice variants disrupting the donor structure (Donor Loss) in the splice donor 5^th^ base (Extended Data Figure 3b), which are not typically considered as pLoF in the current practice. One representative example was rs200831171 – a splice donor 5^th^ base variant of *APOA5* and associated with higher TG concentrations. This intronic variant has high donor loss potential (DS_Donor Loss_ = 0.97) and was associated with increased TG levels with the largest effect size [β = 1.10 (SE 0.079), *P* = 7.0 × 10^−44^] among 6 EWS coding variants in *APOA5* associated with TG (Extended Data Figure 3c). Including this variant, we identified 15 cryptic splice variants with EWS (Supplementary Table 6). Overall, cryptic splicing variants showed equivalent effect sizes with pLoF variants [median β_Cryptic Splice_= 1.092 (IQR 0.601 – 1.118), normalized to pLoF as 1, *P* = 0.71 by Wilcoxon Rank Sum test, Extended Data Figure 3d], and larger effect sizes than missense variants [median β_Missense_ = 0.408 (0.136 – 0.701), *P* = 7.0 × 10^−4^].

## Novel rare variant association outside of established lipid loci

We identified associations for several variants residing outside established lipid loci (Extended Data Figures 4, Supplementary Table 6). One example is a rare missense variant GYS2 p.Y636H (MAF = 0.0431%), which showed significant associations with decreased TC, LDLC, and HDLC [β_TC_ = −0.24, *P*_TC_ *=* 2.3 × 10^−15^; β_LDLC_ = −0.19, *P*_LDLC_ *=* 4.0 × 10^−10^; and β_HDLC_ = −0.22, *P*_HDLC_ *=* 7.1 × 10^−14^]. *GYS2* encodes glycogen synthetase 2, is expressed in the liver^32^, and is a causal gene for glycogen storage diseases^33^. Another example is the *STS* gene on chromosome X. A rare missense variant (p.H439R) in this gene was associated with decreased HDLC. *STS* encodes steroid sulfatase which is directly involved in steroid metabolism^34^. Other novel loci identified by this study include *SH3TC1* (TC), *ETV6* (TC), *PCSK6* (TC), *PCSK9* (HDLC), *POR* (HDLC), and *PTPRB* (TG).

## New insights into causal genes within established lipid loci

Lead variants in genome-wide association studies (GWAS) are typically common and non-coding, and the causal gene is therefore unclear. Rare variant association study more directly interrogates gene product perturbation providing greater confidence in causal gene inference. One such example is 1q21.1, an established HDLC GWAS locus comprising 21 genes (Extended Data Figure 5a). A rare pLoF variant in only *PDZK1* at 1q21.1 was associated with increased HDLC levels (MAF 0.016%, β = 0.30, *P =* 1.1 × 10^−10^), strongly implicating *PDZK1* as the causal gene at this locus. The gene product of *PDZK1* is known to interact with the known HDLC-related gene *SCARB1*^35^. Another example is *SREBF1,* which is a master regulator for lipogenesis^36^. rs114001633 is a rare missense variant in *SREBF1* and associated with higher TC levels (MAF = 0.74%, β = 0.0442, *P =* 2.8 × 10^−9^) and 372kb from the index GWAS non-coding variant (Extended Data Figure 5b). Other examples included a missense association on the androgen receptor (*AR*) p.Q799E in chromosome X with HDLC (Extended Data Figure 5c) and *CREB3L1* in chromosome 11 with TG (Extended Data Figure 5d).

While 58% (87/150) of these putatively effector genes harboring coding variants with EWS were the nearest genes of GWAS lead variants in the loci, the rest (42.0%) were not (Supplementary Table 11). By systemic conditioning analysis and introducing rare-coding alleles as covariates, we confirmed independence of rare-coding associations and common genetic associations (Extended Data Figures 6a and 6b, Supplementary Notes V). Reflecting functional relevance, we observed stronger enrichment of genes harboring rare coding variants with EWS than the nearest genes to the common variant GWAS signals (Extended Data Figures 7a, 7b, and 7c, Supplementary Table 12, Supplementary Notes VI).

## Population enriched coding associations and shared effect sizes

Inclusion of the diverse populations enabled testing for associations with ancestry enriched alleles. By intra-population meta-analysis, we identified 655 signals in European-like (EUR) populations, 124 in African-like (AFR) populations, and 45 in Admix-American-like (AMR) populations (Fig. 3a). Most of these signals are population-specific (631/655 associations were specific for EUR, 105/124 for AFR, and 18/45 for AMR), and overall we identified 130 lipid associated alleles that were only significant in non-EUR populations. These alleles are exclusively or dominantly found in AFR/AMR populations (Fig. 3b).

**Fig. 3.**
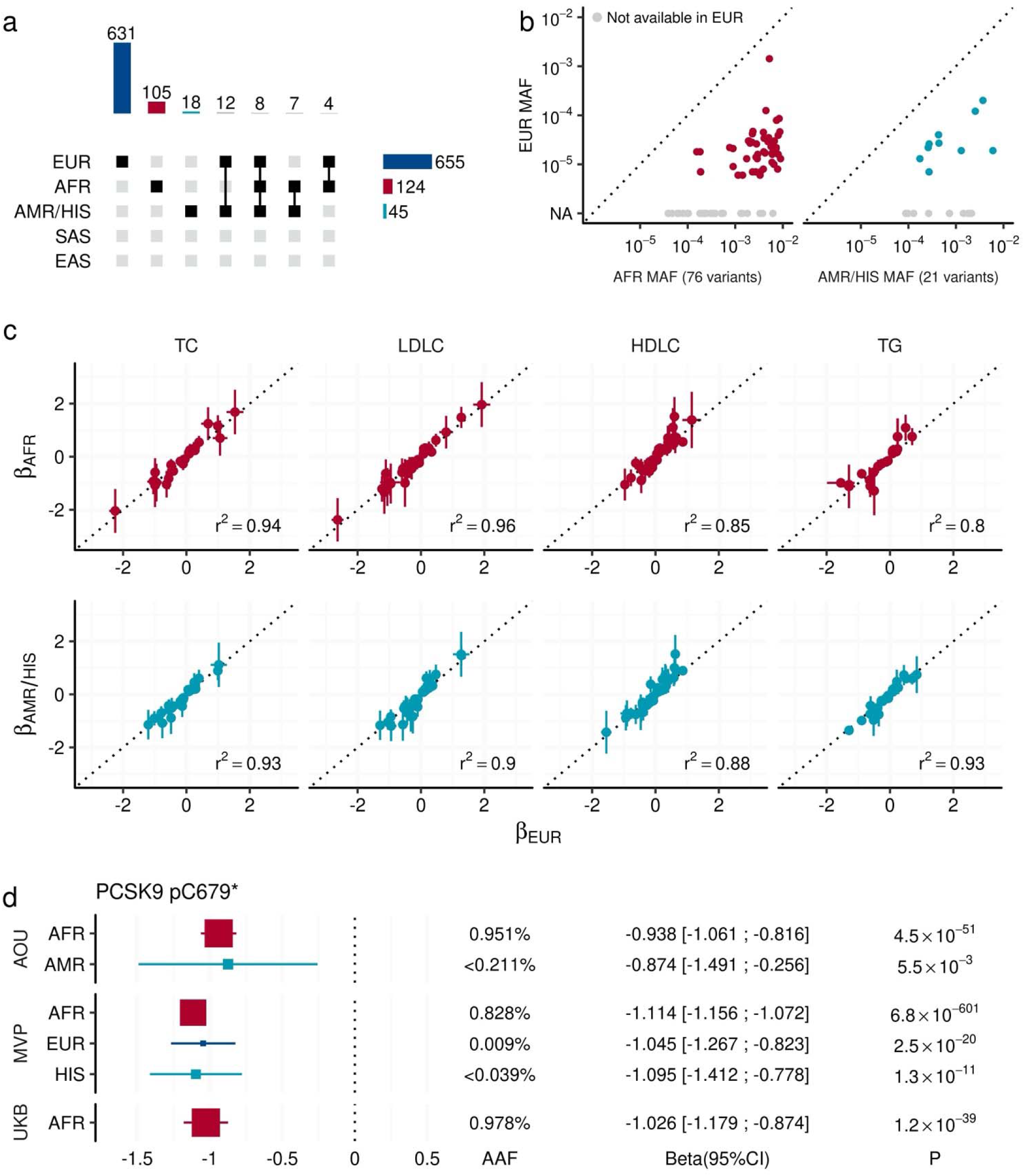
Shared allelic effects across diverse populations. **a.** The upset plot describes the combinations of populations that observed EWS signals through intra-population meta-analysis. The bar chart at the top quantifies the number of EWS associations across various combinations of populations. Each bar represents the total number of associations observed for specific combinations of populations, as indicated by the connected points in the central matrix. The central matrix shows the population combinations involved in each set of associations, where filled squares indicate the populations included in a particular combination. In the right panel, the horizontal bar chart shows the number of associations observed within each population individually. **b.** Allele frequency comparison for non-EUR specific signals. Each point represents an EWS association that is significant only in non-EUR groups (AFR in the left panel and AMR in the right panel). The vertical axes show the minor allele frequency in EUR, while the horizontal axes show the minor allele frequency in AFR or AMR. Gray points indicate variants that were not tested in the EUR group due to low allele frequencies. **c.** Observed effect sizes across studies and populations. Each point indicates variant-trait pair with EWS. The horizontal axis shows the effect sizes in the EUR population. The vertical axes show the effect sizes in AFR and AMR/HIS populations. The error bars represent the 95% confidence interval. R^2^ indicates the squared Pearson’s correlation coefficients of effect sizes. **d.** Consistent effect size of PCSK9 p.C679* (stop gain) variant across multiple populations. The rectangles indicate effect sizes of PCSK9 p.C679* on blood LDLC level in the studied population. The error bars show its 95% confidence interval. The size of rectangles is proportional to AAF. The *P*-values were calculated by linear mixed model with two-sided test. The *P*-values were not adjusted for multiple testing correction. AAF, Alternate Allele Frequency; EWS, Exome Wide Significance; AFR, African-like population; ASN, Asian-like population; AMR, Admixed-American-like population; EAS, East-Asian-like population; EUR, European-like population; HIS, Hispanic-like population; SAS, South-Asian-like population; TC, Total Cholesterol; HDLC, High density lipoprotein cholesterol; LDLC, Low Density Lipoprotein Cholesterol; TG, Triglycerides. MVP, Million Veteran Program; UKB, UK Biobank; AOU, All of Us Research Program.

While we observed significant differences in variant frequencies, we found highly similar effect sizes between genetically dissimilar groups (R^2^ ∼ 0.9, Fig. 3c) for EWS variants. One example is a stop gain variant in *PCSK9* (Fig. 3d), which is dominant in AFR (MAF_AOU-AFR_ = 0.951%, MAF_AOU-AMR_ < 0.211%, MAF_MVP-AFR_ = 0.828%, MAF_MVP-EUR_ = 0.009%, MAF_MVP-HIS_ = 0.036%, MAF_UKB-AFR_ = 0.978), but included consistently large positive effects on HDLC levels [median β = −1.036 (range −1.140 – −0.874)] across populations.

As demonstrated with the polygenic risk score, estimates from large population studies are expected to be valuable resources for assessing individual risk. To explore the feasibility of a rare variant-based risk score, we estimated the carrier frequencies of these alleles in the study populations. The prevalence of lipid-related variants was 67.5% in MVP and 74.0% in UKB overall, but there were significant differences among genetically similar groups, with the highest in EUR (MVP 75.7%, UKB 75.2%) and the lowest in ASN/EAS (MVP 15.4%, UKB 5.5%), likely due to differences in the size of the discovery analysis (Extended Data Figure 8, Supplementary Table 13).

## Contribution of rare coding variants in trait variance

Recent studies suggest additional contribution of the rare variants to the trait variance is not explained by common variants. Using LD-independent rare coding variants with EWS association, we estimated phenotype variance explained (PVE) for each trait. Collectively, rare coding variations contributed to additional 2.03 – 3.75 % of PVE in blood lipids corresponding 15.8 – 22.1% of PVE by common variants (Extended Data Figure 9a, Supplementary Table 14). While per-variant PVEs are slightly lower in rare variants [median (IQR) 0.00645% (0.00392% – 0.0119%)] than common variants [0.00667% (0.00446% – 0.0129%), *P* = 0.002 by Wilcoxon Rank Sum test], sum of the PVE of rare variants showed substantially larger per-variant PVE. The largest PVE by rare coding variants in a single gene was observed in *PCSK9* for LDLC and TC (1.17% and 0.86%, respectively), followed by *APOB* for LDLC and TC (0.97% and 0.65%), *APOC3* for TG and HDLC (0.942% and 0.422%), *LDLR* for LDLC (0.31%). Notably, in 20.6% (39/189) of lead variant - gene pairs, the sum of PVE by rare coding variants exceeded PVE by GWAS leading variant (Extended Data Figures 9c and 9d).

## Insights from recessive modeling

We identified 110 variant-trait pairs with significant associations in the recessive model (*P* < 4.4 × 10^−9^, Fig. 4, Supplementary Table 7). Among these associations, we observed several examples of recessive effect sizes substantially larger than expected from a purely additive model. One example is *ANGPTL4* p.E40K on TG with larger effect sizes in the recessive model (β_Recessive_= −0.845) compared to the additive expectation (2 × β_Additive_ = −0.544). Another example is *TM6SF2* p.L156P which showed > 3 times higher effect size on LDLC in the recessive model [β_Recessive_ = −0.942, *P* = 1.1 × 10^−32^] compared to the additive expectation (2 × β_Additive_ = −0.307). Heterozygosity for this variant has been linked to hepatic triglyceride accumulation and impaired VLDL (a hepatic-precursor of LDL) intracellular trafficking^37^. Another example was observed in *HBB* p. E7V (rs334) – the causal variant for sickle cell anemia^38,39^ and TC or LDLC. While the additive associations were weak for these traits (P_TC_ = 0.0005 and P_LDLC_ = 0.017), the recessive associations showed the largest effect sizes (β_TC-Recessive_= -1.26, *P*_TC-Recessive_ = 2.9 × 10^−19^ and β_LDLC-Recessive_ = −1.12, *P*_LDLC-Recessive_ = 8.3 × 10^−12^) among recessive associations. Strong recessive associations were also observed in *ABHD15* pLoF and lower TG (β_TG-Recessive_= -0.586, *P*_TG-Recessive_ = 5.8 × 10^−11^). In the heterozygote state, the association was not observed (*P* = 0.12). *ABHD15* is known to interact with *PDE3B* and associated with insulin signaling^40^. While recessive inheritance has been emphasized in the context of FH, we did not detect strong recessive associations in the previously suggested recessive genes.

**Fig. 4.**
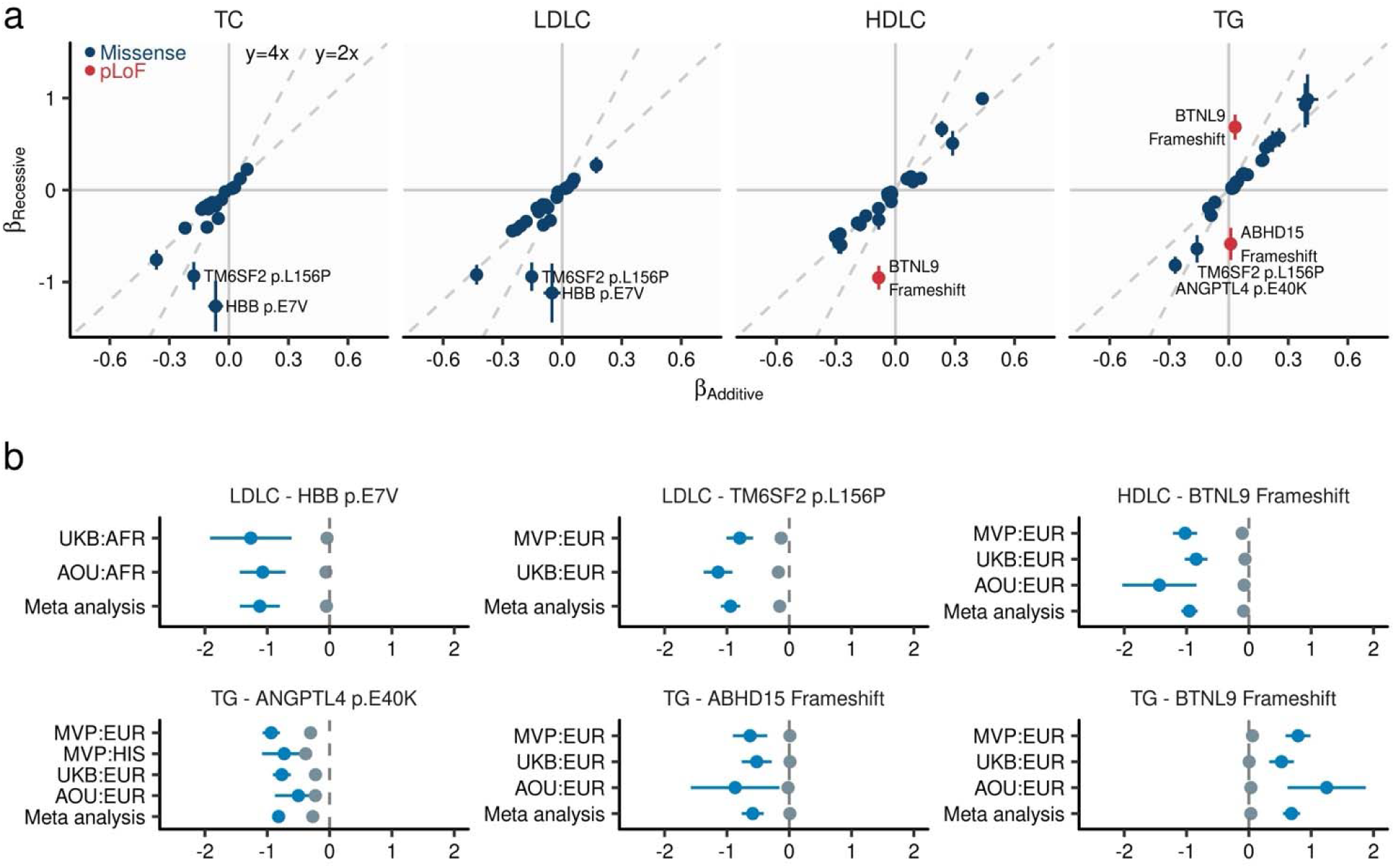
Recessive alleles associated with blood lipids. **a.** Comparison of effect sizes between additive and recessive models. The horizontal axis displays the effect size as estimated by linear mixed model under additive assumption, while the vertical axis shows the effect size estimated under recessive assumption (Methods). Each dot indicates a genetic variant, with the error bar representing the 95% confidence interval. Dashed lines represent the predictions of recessive effect sizes based on the additive model estimates (y = 2x) and estimates that are twice as large (y = 4x) as those from the additive model. **b.** Effect size from population-wise or meta-analysis estimates for variants with the largest deviations in recessive estimates from the predicted effect sizes based on additive model estimates. Gray dots represent additive effect sizes, while dark blue dots correspond to recessive effect sizes calculated by linear mixed model. Error bars indicate 95% confidence intervals. TC, Total Cholesterol; LDLC, Low Density Lipoprotein Cholesterol; HDLC, High Density Lipoprotein Cholesterol; TG, Triglycerides; MVP, Million Veteran Program; UKB, UK Biobank; AFR, African-like population; AMR, Admixed-American-like population; ASN, Asian-like population; EAS, East-Asian-like population; EUR, European-like population; HIS, Hispanic-like population; SAS, South-Asian-like population.

## Pathogenicity reassessment of FH variants

Curated pathogenic variants play a crucial role in the molecular diagnosis of familial hypercholesterolemia (FH). To contribute to this essential resource, we re-evaluated curated variants using our population-scale genomic dataset. By intersecting 6,520 FH-related variants reported in ClinVar database^41^ with 1,601 tested variants in this study, we identified 86 pathogenic/likely pathogenic (P/LP) variants, 268 benign/likely benign (B/LB) variants, and 704 variants of uncertain significance (VUS) in *PCSK9/APOB/LDLR*. The B/LB variants showed a higher allele frequency compared to other classes (Fig. 5a). More than half of the P/LP variants (45 out of 83) are associated with higher LDLC levels (*P <* 0.05/1,601, Fig. 5b) with median β = 1.58 SD_LDLC_ per allele (range 0.51 - 2.61). Importantly, despite fixed clinical categories of pathogenicity, expressivity varied and was overlapping (Fig. 5c).

**Fig. 5.**
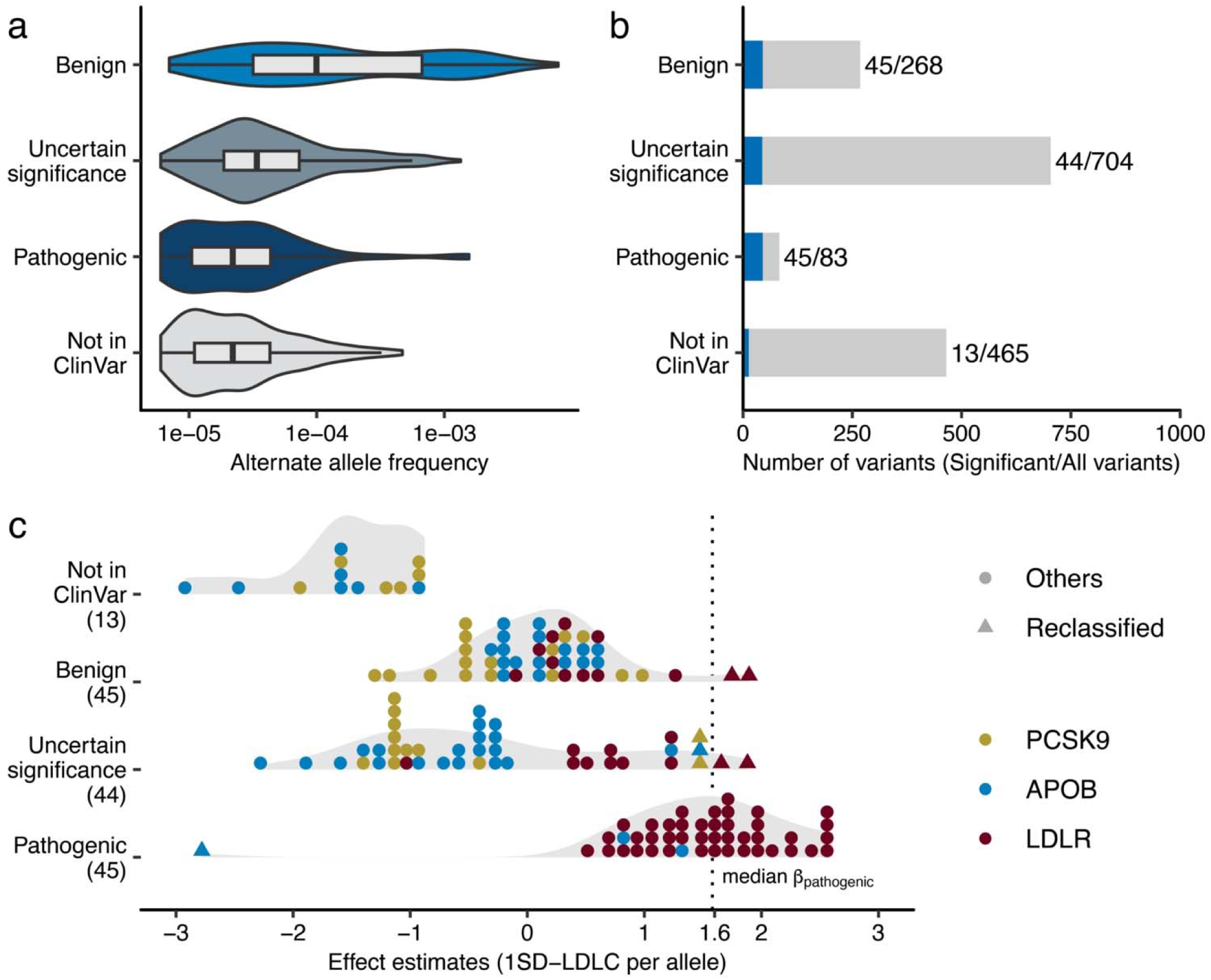
Re-evaluation of clinically curated pathogenic variants for FH. **a.** Variant allele frequencies of FH-related ClinVar variants observed in the study. The rectangles illustrate the interquartile range of the minor allele frequencies, with the bottom and top edges representing the first and third quartiles, respectively. The line inside the rectangle denotes the median and the whiskers extend from the quartiles to the smallest and largest observed values, within a distance no greater than 1.5 times the interquartile range. **b.** Phenotype associations of FH- related ClinVar variants. The height of the bar indicates total number of variants in the category, and the blue color indicates the proportion of the variants significantly associated with clinical LDLC levels in this study. Statistical significance determined using Bonferroni adjustment. **c.** Distribution of the effect sizes for ClinVar FH associated variants determined in this study. Each dot represents a variant in *PCSK9*, *APOB*, or *LDLR*. The color of each dot indicates the associated gene. The dashed, vertical line indicates median effect size for established pathogenic variants. Triangles indicate variants of uncertain significance with large effect sizes, as well as pathogenic variants with a negative effect size on clinical LDLC levels. SD, Standard Deviation; LDLC, Low Density Lipoprotein Cholesterol. FH, Familial Hypercholesterolemia.

We identified eight variants across the B/LB/VUS categories with equivalent effect sizes [median β = 1.66 (range 1.43 - 1.86), Supplementary Table 15] to P/LP variants, including two missense variants in *PCSK9* (p.E40K, p.E197K), one in *APOB* (p.K3524T), and four in *LDLR* (p.H327Y, p.R440G, p.L456P, p.A705P). Among these, *LDLR* p.H327Y is enriched in SAS [MAF = 0.048%, β = 1.75 (SE 0.32), *P* = 5.4 × 10^−8^] but the pathogenicity of this variant was inconclusive in ClinVar. However, its highly significant association with a large effect size on LDLC apart from the median effect size of established P/LP variants supports a pathogenic role of this variant in FH. Another variant, *LDLR* p.L456P, was enriched in AFR [MAF = 0.0066%, β = 1.66 (SE 0.25), *P* = 6.4 × 10^−11^]. We also identified a previously considered pathogenic variant with a negative effect size. A missense variant in *APOB* (p.R490W) showed a strong negative association [β = −2.78 (SE 0.32)] with LDLC levels. This variant is predicted to be a cryptic splice variant with a high DS (DS_Donor Gain_ = 0.98), suggesting it introduces a loss-of-function change in the *APOB* gene and decrease blood LDLC level.

## Clinical outcomes of lipid associated alleles

To connect the lipid related alleles and clinical outcomes, we tested for 800 lipid associated alleles identified in this study with prevalent/incident CAD. We used logistic regression framework to test for significant associations between the lipids associated variants and the occurrence of CAD (Methods). We observed positive associations of TC, LDLC, TG with CAD risk (Fig. 6, Supplementary Table 16) including several strong associations for known FH pathogenic variants in LDLC (p.C197Y. p.C184Y, Splicing variant). On the other hand, HDLC levels were not uniformly associated with CAD risk, as exemplified by the known association between higher HDLC level and increase CAD risk by *SCARB1*. Several established lipid related genes (*PCSK9*, *APOB, NPC1L1*, *ANGPTL3/4*, *APOC3*, and *LDLR*) were associated with lower LDLC/TG and decreased CAD risk with nominal significance (*P*_CAD_ < 0.05). Overall, we identified five genes significantly associated with CAD (FDR_CAD_ < 0.05) including *RORC*, *CFAP65*, *GTF2E2*, *PLCB3*, and *ZNF117*. Among these genes, *RORC* has high potential as a new therapeutic target to prevent CAD. In vitro and in vivo studies suggested beneficial effect of silencing *RORC* in the development of atherosclerotic disease^42,43^ consistent with protective effect of pLoF in *RORC* for CAD observed in this study.

**Fig. 6.**
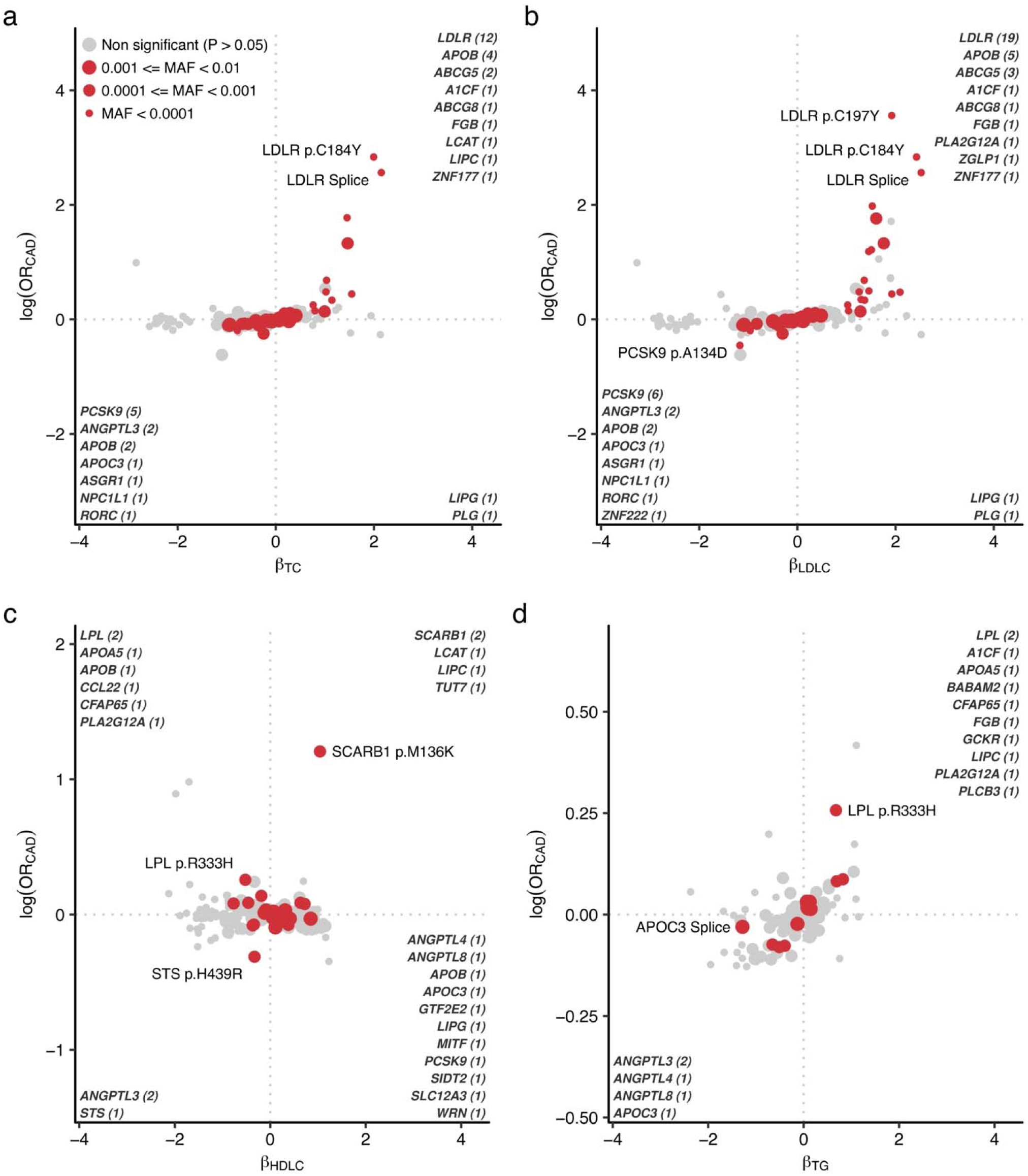
CAD risks in blood lipid associated alleles. Scatter plots indicate effect size in lipids on the horizontal axes and log odds ratio for CAD on the vertical axes. Nominally associated (*P* < 0.05) variants with CAD were highlighted in red and the sizes of the points indicating minor allele frequency. The associated gene names are highlighted in the corner of quadrant and the number of associations were indicated. CAD, coronary artery disease; OR, odds ratio; MAF, minor allele frequency; TC, total cholesterol; LDLC, low density lipoprotein cholesterol; HDLC, high density lipoprotein cholesterol; TG, triglycerides.

In this study, we conducted the largest rare variant association study for blood lipids to-date. The substantial sample size enabled the analysis of single rare variants as opposed to more conventional aggregation of rare variants into a statistical unit for burden testing. This analysis not only advances novel mechanistic insights but also improves the clinical interpretation of Mendelian dyslipidemia genotypes beyond the current clinical classification schema. Overall, this study demonstrated the capability of population-based analyses of to identify rare coding alleles with both mechanistic and clinical implications.

Importantly, our study expands allelic diversity by including large cohorts from non-European-like populations resulting in the discovery of 130 alleles that are exclusively or dominantly observed in the non-European-like populations. alleles, We typically observed consistent and highly similar effect sizes across populations despite differences in allele frequencies. The transferability of associated rare coding alleles may reflect the causality of these alleles and consistent with our observations from the systematic evaluation of rare variant burden testing across various traits^44^.

In addition to insights specific to blood lipids, our study provides several observations that may be generalizable. Specifically, our expansive rare variant association study in highly heritable phenotypes allowed qualitative/quantitative assessment of variant characteristics behind significant associations. In our study, associated variants are significantly enriched in functional variant classes (high confidence pLoF or deleterious missense variants) highlighting the importance of further effort for precise classification of variant functionality. We further implemented machine learning-based splice site prediction^25^, and successfully re-classified previously underestimated variant class. The cryptic splice variants showed constraint pattern and enrichment by associated variants equivalent to canonical pLoF variants.

The broad range of allele-specific analyses also allowed us to infer the variant effects on gene function. First, we observed highly concordant effect direction of pLoF alleles (> 99%) on the same gene proxied by phenotypic expression confirming the findings from previous study^45^. Also, most (87%) missense variants showed concordant effect directions with pLoFs, however, the remainder (13%) had opposite effects, indicative of hypermorphic characteristics. *In silico* deleterious prediction is not effective to capture these hypermorphic alleles and these variants might be missed by variant filters for gene-based testing despite their empiric functional significance. Increasingly available large-scale genetic analysis across diverse phenotypes and populations, focusing on rare coding variations, may expand the list of hypermorphic alleles and inform models to better detect this phenomenon across genes and domains.

We also conducted recessive modeling and identified multiple strong associations. Intriguingly, the recessive associations are robustly shared across studies and populations. Aligned with previous studies focusing on binary traits^46^, some rare alleles have a prominent recessive effect not captured by standard additive modeling, suggesting a contribution to the missing heritability.

Additionally, using estimated effect size and statistical significance driven by population scale association analysis, we re-assessed a curated database considered a gold standard for clinical genetic diagnosis. We confirmed the accuracy of most variant annotations aligned with previous study^47^ and further provided evidence toward potential re-classification of the pathogenicity of other variants. Especially, we found two non-European enriched candidate variants, aligned with previous studies. including ours,^19,48,49^ that reported under diagnosis of genetic disease in the non-European population. Importantly, we observed a range of expressivity for pathogenic alleles that was associated with clinical outcomes.

In conclusion, we conducted a rare variant focused genetic study for blood lipids involving over a million individuals, yielding hundreds of rare alleles associated with blood lipids and improved mechanistic understanding of rare variant associations. Our study suggests that population-scale rare variant analysis is now adequately powered for heritable phenotypes, allowing for the classification of rare pathogenic alleles and providing new insights into variant expressivity/penetrance, toward improved diagnosis and more quantitative prognosis.

## Online Methods

### Ethics oversight

This study received ethics approval from the Veterans Affairs Central Institutional Review Board under Institutional Review Board protocol number 16-06. The study protocols were approved under protocol numbers 2016P002395 and 2021P002228 by the Mass General Brigham Institutional Review Board. The analysis for UKB was performed under application number 7089.

### Blood Lipids Phenotyping

The UK Biobank (UKB) is a volunteer cohort of approximately 500,000 residents aged 40 to 69 years of age living in the United Kingdom recruited 2006 - 2010^21^. In the UKB, blood lipids were measured using blood samples collected at the enrollment. We adjusted total cholesterol (TC) and low density lipoprotein cholesterol (LDLC) levels by dividing 0.7 for individuals prescribed lipid-lowering medication at the enrolment as previously described^50^.

The Million Veteran Program (MVP) is a national hospital-based cohort initiated in 2011 by the United States Department of Veterans Affairs (VA). Recruitment was conducted in the VA affiliated hospitals across the United States^20,51^. In the MVP, lipid phenotypes were derived from longitudinal lipid measurements over the time. For TC, LDLC and triglycerides (TG), we utilized the highest value recorded, while for high density lipoprotein cholesterol (HDLC), we selected the lowest value as previously described^52^.

The All of Us Research Program is a U.S.-based population cohort that began enrollment in 2018 under the National Institutes of Health (NIH). Participants were enrolled through a network of more than 340 recruitment sites. In AOU, lipid phenotypes were derived similarly to those in MVP. To minimize potential overlap between AOU and MVP participants, we excluded AOU participants who answered the baseline survey, indicating they are receiving healthcare from the Veterans Affairs (n = 13,400), from the analysis.

### CAD Phenotyping

In the UKB and AOU, we ascertained CAD cases based on at least one of the following criteria: a) any ICD code in the in-hospital record or death registry (I21 – I25 in ICD10; 410 - 414 in ICD9), or b) any procedure code for coronary revascularization (K40 – K45, K49, K50, and K75 in OPCS4, 33510 – 33523, 33533 – 33536, 92920 – 92950 in CPT4). In the MVP, we used a previously established CAD definition^53^. ICD9, ICD10, and CPT codes along with self-report were used to determine CAD cases and controls. Qualifying codes were those pertaining to acute myocardial infarction (inpatient only), stable ischemic heart disease (inpatient or outpatient), and coronary revascularization (inpatient and outpatient). Cases were individuals who had at least 2 qualifying codes on different dates within a 12-month period. Controls were individuals who carried no codes and who did not self-report a history of coronary artery disease.

### Quality control for microarray genotyping in UKB

We conducted sample quality control as follows. Among 488,175 individuals, we removed samples with aneuploidy (N = 651), sex-gender mismatch implying phenotypic quality issues (N = 378), higher heterozygosity or missing outlier (N = 739) leading to a total of 1811 (0.4%) samples removed. 486,364 quality control passed individuals remained (9,454 AFR, 2,413 AMR, 2,582 EAS, 461,352 EUR, 10,563 SAS).

### Population ascertainment in UKB

Using reference population data from 1000 genomes project and microarray genotypes in UKB using the Affymetrix UK BiLEVE Axiom and UK Biobank Axiom arrays, we determined genetically determined population ascertainment. First, we extracted quality-controlled variants from 1000 genomes data [non-palindromic single nucleotide variant (SNV), minor allele frequency (MAF) > 1%, a population specific Hardy Weinberg equilibrium *P*-value > 1 × 10^−6^]. Next, we extracted the intersection of the quality controlled 1000 genomes data and the study population. Using intersected variants, we pruned variants on the 1000 genomes data using PLINK2^54^ software (Jun 3, 2022, release) with --indep-pairwise option (window size 50, sliding window size 10, R^2^ < 0.2). Which yields 224,993 variants. Using pruned variants, we calculated the SNV weights for genetic principal components. Then we projected study participants to the principal components space. Using the 1000 genomes reference population annotation, we trained the k-nearest neighbor model using class R package (version 7.3). Then we split study cohort into the five genetic populations (African-like, AFR; Admixed-American-like, AMR; East-Asian-like, EAS; European-like, EUR; and South-Asian-like, SAS) and conducted association analyses separately to minimize potential effects of heterogeneity.

### Quality control for WES in UKB

For genotype level quality control, first, we utilized Hail’s^55^ ‘split_multi_hts’ function to divide multiallelic sites. We then filtered out low-quality genotypes based on the following criteria: i) Genotyping quality less than or equal to 20. ii) Genotype depth (DP) either less than or equal to 10 or greater than 200. iii) For heterozygous genotypes: (DP_Reference_ + DP_Alternate_)/(DP_Total_) > 0.9 and DP_Alternate_/DP_Total_ > 0.2. iv) For alternate homozygous genotypes: DP_Alternate_/DP_Total_ > 0.9.

These processes retained 26,645,535 variants in the 454,756 sequenced samples. We excluded i) 6,131,710 variants due to high missingness (missing rate > 10%). ii) 47,441 variants that deviated from the Hardy-Weinberg equilibrium (*P*_Hardy-Weinberg equilibrium_ < 1 × 10^−15^). iii) 364,207 variants located within low-complexity regions^56^. Cumulatively, we excluded 6,289,813 variants, resulting in 20,355,722 retained variants.

Among 454,756 individuals whole exome sequenced (450K UKB release using Deep Variant^57^ for variant detection), we identified 452,929 individuals overlapping array genotyped data. For these individuals, we conducted sample-level quality control. First, we calculated array-exome genotype discordance rate and F-statistics in non-pseudo autosomal region X-chromosome variants to detect potential sample or phenotypic swapping in exome data. For this analysis, we used pruned and stringent variant quality control criteria (missingness < 1%, MAF > 0.1%). We identified 27 potential sex- swapping (27 Females with F-statistics > 0.6, 0 Males with F-statistics < 0.6) and 0 discordant genotypes between exome and array data (non-reference homozygote concordance rate < 0.8). We calculated array-exome discordance using pruned, non-palindromic, high-quality exome data and the corresponding array data. Next, we removed samples with a high missing rate (> 10%, N = 12). Then, we filtered samples using the following autosomal quality control outlier metrics [outside mean ± 8 standard deviation (SD)]: heterozygous/homozygous rate (N = 761), transition/transversion rate (N = 0), SNV/Insertion-Deletion ratio (N = 2), number of singletons (N = 283). In total, 1,052 (0.2%) samples were removed, resulting in 451,877 samples in the final dataset. After removing these samples, we also removed 226,083 monomorphic variants in the dataset retaining 20,129,639 variants among the 451,877 samples in the final dataset.

### Quality control for microarray genotyping, population ascertainment, and imputation in MVP

Genotyping was performed using the custom Axiom array (MVP1.0), and variant and sample quality control was described in detail previously^20^. We used the latest release (release 4) data for this analysis. Release 4 data included array-genotypes and genetic dosage imputed to the TOPMed imputation r2 reference panel^22^. The quality-controlled sample size was 657,242. We grouped participants into four population groups [AFR, Asian-like (ASN), EUR, and Hispanic-like, HIS] following the harmonized ancestry and race/ethnicity (HARE) algorithm previously established in MVP^58^.

### Quality control for WGS in MVP

To confirm the accuracy, sensitivity, and specificity of the imputation, we have utilized the initial release of whole genome sequencing data in the MVP study. This data was collected and sequenced with a focus on elucidating the pathophysiology of COVID-19 infection from their genomes. The sequencing was performed using Illumina’s Sequencing by Synthesis technology to a targeted depth of 30x. Individual variant calling from 10,413 samples was performed on the cloud-based data and task management framework Trellis^59^. In summary, reads were aligned with BWA-MEM (version 0.7.15) on the GRCh38 reference genome, and variant calling was performed in GATK 4.1.0.0 using the haplotypeCaller function. Genotypes of all samples were aggregated into a matrix table using gVCF Combiner implemented in Hail^55^ for additional quality-control steps. In summary, we retained high-quality genotypes by applying the following steps: I. Variants in low complexity regions and ENCODE blacklist regions were removed. II. Variants within regions of atypical sequencing depth (DP < 10 or DP > 400) were discarded. For haploid genotypes on sex chromosomes, a minimum DP > 5 was required. III. Genotypes were retained if sites were: a. Homozygous reference with Genotype Quality > 20, or b. Alternate homozygotes with Phred-scaled likelihood of the genotype for reference homozygotes (PL[0]) > 20, and the ratio of depth for alternate alleles (DP_ALT_) to total depth at the site (DP_ALT_/DP_SITE_) > 0.9, or, c. Heterozygous with PL[0] > 20, and the ratio of the sum of DP_ALT_ and depth for reference alleles (DP_REF_) to DP_SITE_ [(DP_ALT_ + DP_REF_)/DP_SITE_] > 0.9, and DP_ALT_/DP_SITE_ > 0.2. III. Variants with high missing rate (> 0.8) and population wide *P*_Hardy-Weinberg equilibrium_ ≤ 1 × 10^−5^ for variants with minor allele frequency (MAF) ≥ 1%, and *P*_Hardy-Weinberg equilibrium_ ≤ 1 × 10^−6^ for variants with MAF < 1% were discarded. IV. Samples with low call rate (≤ 0.97) or low overall sequencing coverage (mean depth ≤ 18) were excluded. This processing resulted in 187,790,701 variants in 10,390 individuals.

### Quality control for WGS in AOU

We curated genotypes from the jointly called WGS call set (version 7) provided AOU^24^. We split multiallelic site to biallelic variants using hail’s split_multi_hts function, then, low quality genotypes flagged as FAIL in FT field was set as missing. The genotypes were export as bgen files and converted to pgen files for quality control procedure and downstream analysis. We filtered variants i) flagged in the FILTER column in original VDS, ii) located in the low complexity region, iii) low call rate (< 90%), iv) monomorphic, v) population specific Hardy Weinberg equilibrium P-value < 1 × 10^−15^. Finally, we excluded flagged individuals (n = 549) and genotype missing rate more than 1% (n = 396).

### Exome-wide association analysis

We utilized the association analysis framework implemented in Regenie software (version 3.1.3)^60^. We used array-based autosomal genotypes for step 1 excluding variants with MAF < 1%, *P*_Hardy-Weinberg equilibrium_ < 1 × 10^−15^, call rate < 98%, and located in the Major Histocompatibility Complex region (Chromosome 6 23 - 37 megabases). We pruned variants using PLINK2^54^ software (Jun 3, 2022, release) with --indep-pairwise option (window size 1000, sliding window size 100, R^2^ < 0.9) by genetic population in each cohort. The association model was adjusted by age, age^2^, sex, and the first ten genetic principal components and blood lipids measurements were inverse rank normalized. For CAD analysis, we used firth logistic regression implemented in Regenie software. We tested all quality-controlled genotypes in exome sequence data in UKB. For the MVP whole genome imputed dataset and AOU WGS dataset, we restricted the analysis to the exome sequence targeting file used for UKB exome sequencing (https://biobank.ndph.ox.ac.uk/ukb/refer.cgi?id=3801) with 50bp flanking in both sides of the target region. After generating summary statistics for each cohort (MVP, AOU, and UKB) and each population (AFR, HIS, ASN, EUR in MVP and AFR, AMR, EAS, EUR, SAS in UKB and AOU), we meta-analyzed the results using the GWAMA^61^ software (version 2.2.2).

The obtained summary statistics were meta-analyzed using GWAMA^61^ fixed effect model. We computed effect size and *P*-values all the variants in exome region irrespective to the variant annotation. While we used summary statistics for synonymous/non-coding variants as reference to contextualize coding associations, the statistical significance of these variants was not considered throughout study. In this study, we primarily applied additive model for the association analysis. In addition to the additive model, we performed association analyses modeling recessive effects. For the additive model, we restricted the analysis for the variants with 5 ≤ MAC and MAF_POPMAX_ < 1% before meta-analysis. For recessive model, we also restricted the analysis for the variants with 5 ≤ estimated minor homozygote counts (number of participants × MAF^2^) and estimated minor homozygote frequency (MAF^2^) < 1%. We reported unadjusted *P*-values without correction for multiple testing throughout the manuscript.

Per the reporting guidelines by the MVP, we have masked the MAF of variants with a MAC of less than equal to 12 in the summary statistics. This measure is implemented to prevent the potential identification of individuals participating in the study. The directions of the effects in the table and summary statistics indicate the variant effect in the alphabetical order: AOU_AFR_, AOU_AMR_, AOU_EAS_, AOU_EUR_, AOU_SAS_, MVP_AFR_, MVP_ASN_, MVP_EUR_, MVP_HIS_, UKB_AFR_, UKB_AMR_, UKB_EAS_, UKB_EUR_, and UKB_SAS_.

### Variant annotation

We utilized a single transcript for each gene based on Gencode v41^62^ canonical and coding transcript (coding transcript set, n = 19,603, Supplementary Dataset) for all annotations. First, we annotated tested variants (variants within ± 50 bases from target region in the UKB exome) with the VEP^28^ software (version 107, aligned with Gencode v41) and selected annotations on the coding transcript set. If we found variants overlapping in more than two transcripts in the coding transcripts set, we selected higher functional consequence. If the consequences were equivalent, we selected the annotation on the longest transcript. We selected predicted loss of function (pLoF, IMPACT HIGH) or missense (IMPACT MODERATE) variants as coding variants. Next, we ran Splice AI^25^ for all the variant tested for the coding transcript set with default parameters. Splice AI returns DS which represent potential for cryptic splicing (Supplementary Notes III). We treated non-pLoF variants with DS > 0.8 as cryptic splice variants and reclassified them as pLoF.

### Missense Score

For further classification of missense variants, we applied ensemble prediction using 29 in-silico prediction models to assess the deleteriousness of missense single nucleotide variants. Using pre-computed in-silico predictions in the dbNSFP^29^ database (version 4.2), we annotated all the missense variants using the dbNSFP plugin for VEP. We binarized the predictions into ’Deleterious’ or ’Tolerant’ using an algorithm-specific threshold. We computed the Missense Score, ranging from 0 to 1, is calculated by dividing the number of Deleterious predictions by the total number of available algorithms for the corresponding variant.

### Pathway enrichment analysis

We compared pathway enrichments between genes identified by rare coding variant associations in this study (1) and those identified by common variant association studies (2). The analysis was restricted to genes included in the coding transcript set (n = 19,603). For genes supported by rare variants (1), we chose those with the smallest P-value within their respective loci. For genes supported by common variants (2), we selected the closest gene to the lead variant identified in a recent large-scale genome wide association study (GWAS) by Graham et al., as reported in *Nature* in 2021^50^. For the selected gene sets, enrichment was tested using the enrichR (version 3.0) R package^63^, considering the following pathway sets: Reactome_2022; KEGG_2021_Human; GO_Biological_Process_2023; GO_Cellular_Component_2023; GO_Molecular_Function_2023; ChEA_2022; ENCODE_TF_ChIP-seq_2015; ENCODE and ChEA Consensus TFs from ChIP-X; and Enrichr Submissions TF-Gene Cooccurrence. Enrichment was considered significant if the P-value was lower than the threshold adjusted by the Holm method.

### Replication analysis

For replication, we utilized data from a previous large-scale exome array study by Lu et al., as reported in *Nat Genet* in 2017^64^. All variants were updated to the hg38 reference genome using the LiftoverVcf function in GATK^65^. We then combined the updated summary statistics from the previous study with those from our current study. In total, we identified 387 combinations of variants and phenotypes that matched between both studies. The concordance of effect sizes and statistical significance was assessed. A directional concordance was noted if the effect direction was the same in both the replication dataset and our study. Statistical significance was defined by a *P*-value in the replication dataset that was smaller than the Bonferroni-adjusted threshold (*P* < 0.05/387).

### Variance Explained

Per variant explained variance (*Var*) was computed by the following formula^50^:

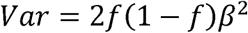

We calculated the variance explained by common variants using the index variant from the latest GWAS for common variants (511 variants for TC, 442 variants for LDLC, 562 variants for HDLC, 480 variants for TG)^50^. To eliminate linked variants in exome wide significant (EWS) rare variants, we employed the *clump* function in PLINK1.9^54^. With the MVP imputed genotypes and UKB WES, we clumped EWS variants using an R^2^ threshold of 0.01. By this process, variants in linkage disequilibrium (R^2^ ≥ 0.01) with any variants that had smaller P-values were excluded. Additionally, EWS variants do not present in the MVP or UKB were omitted from the analysis. As a result, 172, 195, 182, and 121 variants from MVP and 179, 197, 185, and 128 variants from UKB were retained for TC, LDLC, HDLC, and TG, respectively.

### Conditioning analysis

To evaluate the independence of genetic signals derived from rare coding variants and common variants, we executed a conditional analysis where rare coding variants were incorporated as covariates. This analysis was performed in addition to using the standard covariates applied in our primary analyses, which included sex, age, the age^2^, and the first ten genetic PCs. For conditioning purpose, we utilized genotype data for rare coding variants with EWS as covariates. In the MVP, we introduced 185, 207, 203, and 131 rare coding variants as covariates for TC, LDLC, HDLC, and TG, respectively. Similarly, in the UKB, 197, 224, 209, and 140 variants were introduced to the model for TC, LDLC, HDLC, and TG, respectively. By comparing the β and *P*-values obtained from the analyses conducted with and without these genotype covariates, we aimed to ascertain the extent to which signals from rare variants are dependent on or independent from those associated with common variants. This approach leveraged the *condition* function available in the Regenie software package. Conditioning was done in both Step 1 and Step 2.

### Pathogenic variant reclassification

We curated pathogenic alleles for Familial Hypercholesterolemia, a well-known monogenic condition linked to severe hypercholesterolemia and premature coronary artery disease, from the ClinVar^41^ database, downloading bulk data on August 16, 2022. We first extracted genetic regions corresponding to the *PCSK9*, *APOB*, and *LDLR* genes from the VCF file. Using the same pipeline as for the tested variants, we annotated these variants and excluded pLoF variants for *PCSK9* and *APOB* due to their known reduction of LDLC levels. We then classified variants as necessary with conflicting interpretations by majority vote. We calculated the difference in evidence [Number of Pathogenic + Likely Pathogenic − (Benign + Likely Benign + Uncertain Significance)]; if the score was greater than 0, the variants were considered Pathogenic/Likely pathogenic; otherwise, they were considered Benign. This process resulted in a categorized list of variants in three classes, i) Pathogenic/Likely Pathogenic (P/LP), ii) Uncertain Significance (VUS), and iii) Benign/Likely Benign (B/LB). We intersected these variants with those in *APOB/LDLR/PCSK9* tested in this study and defined the pathogenic effect size by taking the median of positive effect sizes from known pathogenic variants. To identify a subset of VUS to be reclassified as P/LP, we ranked the variants by their effect sizes and grouped them, accordingly, ensuring that the median effect size of this group was larger than that of the known P/LP variants.

### External data

For replication, we obtained summary statistics from previous exome array-based study (Lu et. al. *Nat Genet* 2017)^26^. We lifted summary statistics from hg19 coordinate to hg38 using LiftOverVcf function in picard software. We removed insertions/deletions due to ambiguousness of alleles (n = 24) and failed in lifting (n = 71). In total, we successfully lifted > 99.96% (292,322/292,417) of variants in the data. For common variant integration analysis, we obtained summary statistics from the latest GWAS (Graham et. al. *Nature* 2021)^50^. We utilized summary statistics from trans population meta-analysis (with_BF_meta-analysis_AFR_EAS_EUR_HIS_SAS_*_INV_ALL_with_N_1.gz, for autosomes and meta-analysis_chrX_AFR_EAS_EUR_HIS_SAS_*_INV_ALL_with_N_1.gz for X chromosome). All summary statistics were downloaded from the Global Lipids Genetics Consortium website (http://www.lipidgenetics.org). For those summary statistics, we successfully lifted more than 99.82% of variants.

## Data availability

Full summary statistics will be publically available after the acceptance of the manuscript through dbGAP. The individual data for AOU, MVP, and UKB is available upon application to the respective organizations. The analysis codes and supplemental data are available at Zenodo (https://zenodo.org/doi/10.5281/zenodo.11092802). The docker/singularity images used in the analysis are publically available through docker hub (https://hub.docker.com/u/skoyamamd).

## Acknowledgments

This research is based on data from the Million Veteran Program, Office of Research and Development, Veterans Health Administration, and was supported by award #BX004821. This publication does not represent the views of the Department of Veteran Affairs or the United States Government. The analysis of UK Biobank was performed under the application number 7089. S.K. is supported by Japan Society for the Promotion of Science (202160643), Uehara Memorial Foundation, and National Heart Lung and Blood Institute (NHLBI, K99HL169733). Z.Y. is supported by National Human Genome Research Institute (K99HG012956). S.H.C is supported by NHLBI (R01HL127564). S.J.J. is supported by the Dutch Heart Foundation (grant no. 03-007-2022-0035). M.S.S is supported by TOPMed (2022-6842.02), D.K. is supported by the Department of Veterans Affairs (VA, IK2BX005759-01), the American Heart Association (DOI: https://doi.org/10.58275/AHA.23SCEFIA1153369.pc.gr.173943), and the Baszucki Research Initiative provided to Stanford Vascular Surgery. This work was supported in part via funding from VA Merit Award I01 BX003362 (K.M.C., P.S.T.) from the VA Office of R&D. P.N. and G.M.P are supported by NHLBI (R01HL142711, R01HL127564).

## Author contributions

S.K., P.T.E., Y.V.S., P.W.W, P.N. conceptualized this project. S.K., Z.Y., D.K., J.E.H., K.C. curated phenotype data. S.K., S.H.C, S.J.J., M.S.S., D.K., J.E.H., J.S.D., P.S.T. curated genotype data. S.K., Z.Y., S.H.C, S.J.J., M.S.S., M.N.T., A.R. analyzed data. S.K., J.E.H., M.N.T., A.R., J.S.D., C.S., I.S., S.M.D., K.M.C., T.L.A., D.J.R., G.M.P., P.T.E., Y.V.S., P.W.W, P.N. interpreted data. S.K., Y.V.S., P.W.W, P.N. prepared the initial draft S.K., Z.Y., S.J.J., M.S.S., D.K., J.S.D., C.S., I.S., S.M.D., K.M.C., T.L.A., D.J.R., G.M.P., P.T.E., Y.V.S., P.W.W, P.N. provided critical review and edits for the manuscript. W.H., P.S.T., K.C., P.T.E., Y.V.S., P.W.W, P.N. supervised the project. A.B., K.L., W.H., P.S.T., K.C., P.T.E., Y.V.S., P.W.W managed the project administration. K.C., P.T.E., Y.V.S., P.W.W, P.N. obtained funding for the project.

## Extended Data Figures

**Extended Data Figure 1.**
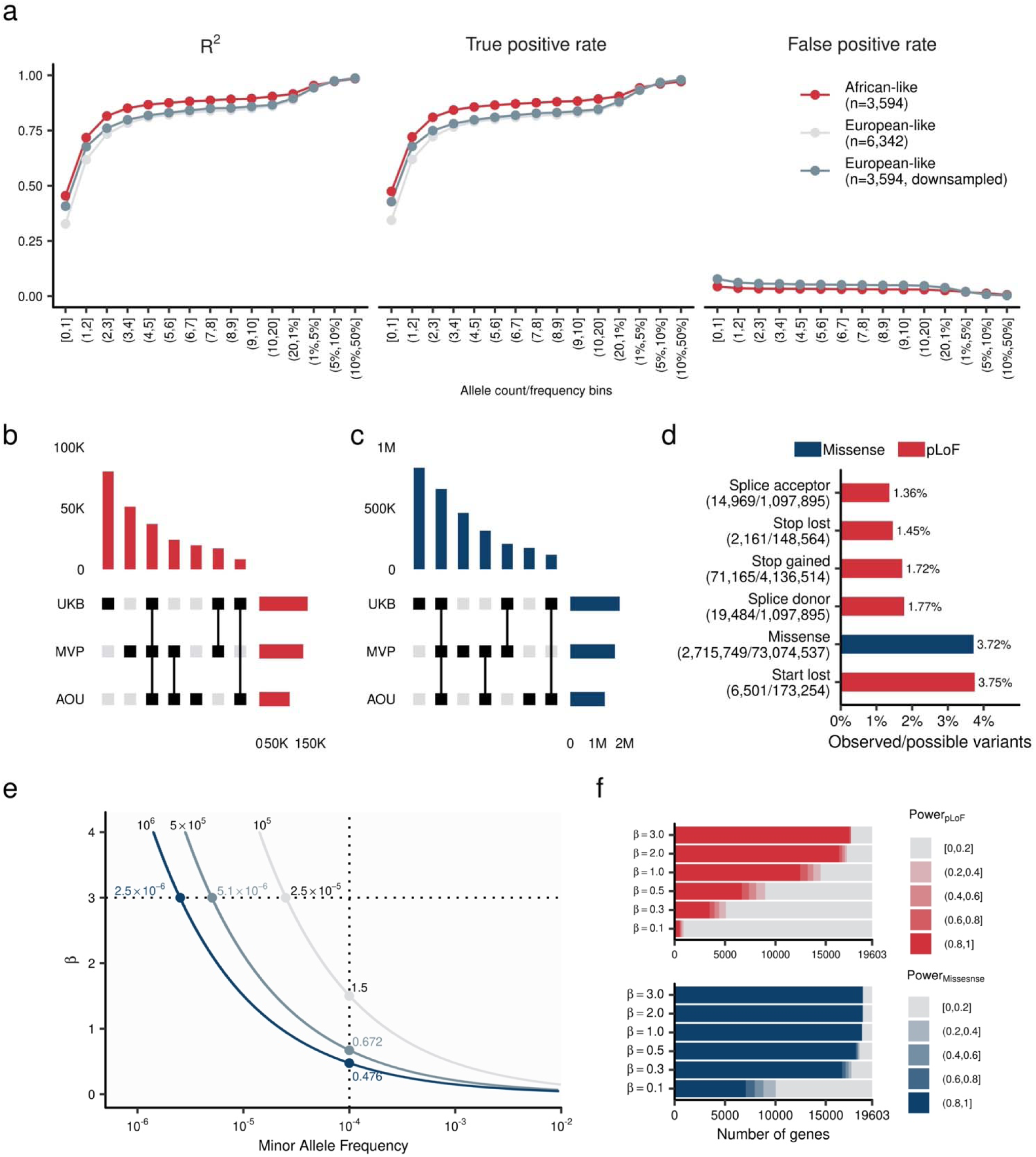
The imputation quality, allelic diversity, coverage, and power in the study. **a.** Imputation accuracy in MVP whole genome imputation date by TOPMed imputation reference panel. Each dot indicates mean R^2^ (Squared Pearson’s correlation coefficient), TPR, and FPR by population and MAC/MAF bins. TPR and FPR were computed by comparing dichotomized hard-called dosage (imputed data) and dichotomized sequenced genotype (WGS data, Supplementary Notes I). **b** and **c.** Shared and unique variants across MVP, UKB, and AOU for pLoF (b) and missense (c) variants. The central matrices define the variant sharing status between MVP, UKB, and AOU. The top panel quantifies the variants within the groups defined in the central matrices. The right panel summarizes the count of variants in each study. **d.** Variant coverage. The relative proportions of SNVs identified in this study is shown as a fraction of all possible SNVs within the target transcripts. **f.** Simulated power curves for different sample sizes. The horizontal axis indicates minor allele frequency, and the vertical axis indicates effect size. The dark blue line indicates 80% power curve at 1 million sample size, the intermediate curve indicates 500K sample size, and the gray curve indicates 100K sample size, respectively. **e.** Power curve for tested genes in this study. The curves indicate most powered pLoF/missense variants in each gene estimated by simulated effect size (β) and observed allele frequency. The color intensity corresponds with β. **f.** Gene based power estimation. The color of the bar charts indicates the highest power of the coding variant in the gene. The top panel shows pLoF variants and the bottom panel shows missense variants. β indicate simulated effect size. TPR, True Positive Rate; FPR, False Positive Rate; MAC, Minor Allele Count; MAF, Minor Allele Frequency; WGS, Whole Genome Sequence; MVP, Million Veteran Program; UKB, UK Biobank; TOPMed, Trans-Omics for Precision Medicine; AFR, African-like population; AMR, Admixed-American-like population; ASN, Asian-like population; EAS, East-Asian-like population; EUR, European-like population; HIS, Hispanic-like population; SAS, South-Asian-like population. pLoF, predicted Loss of Function.

**Extended Data Figure 2.**
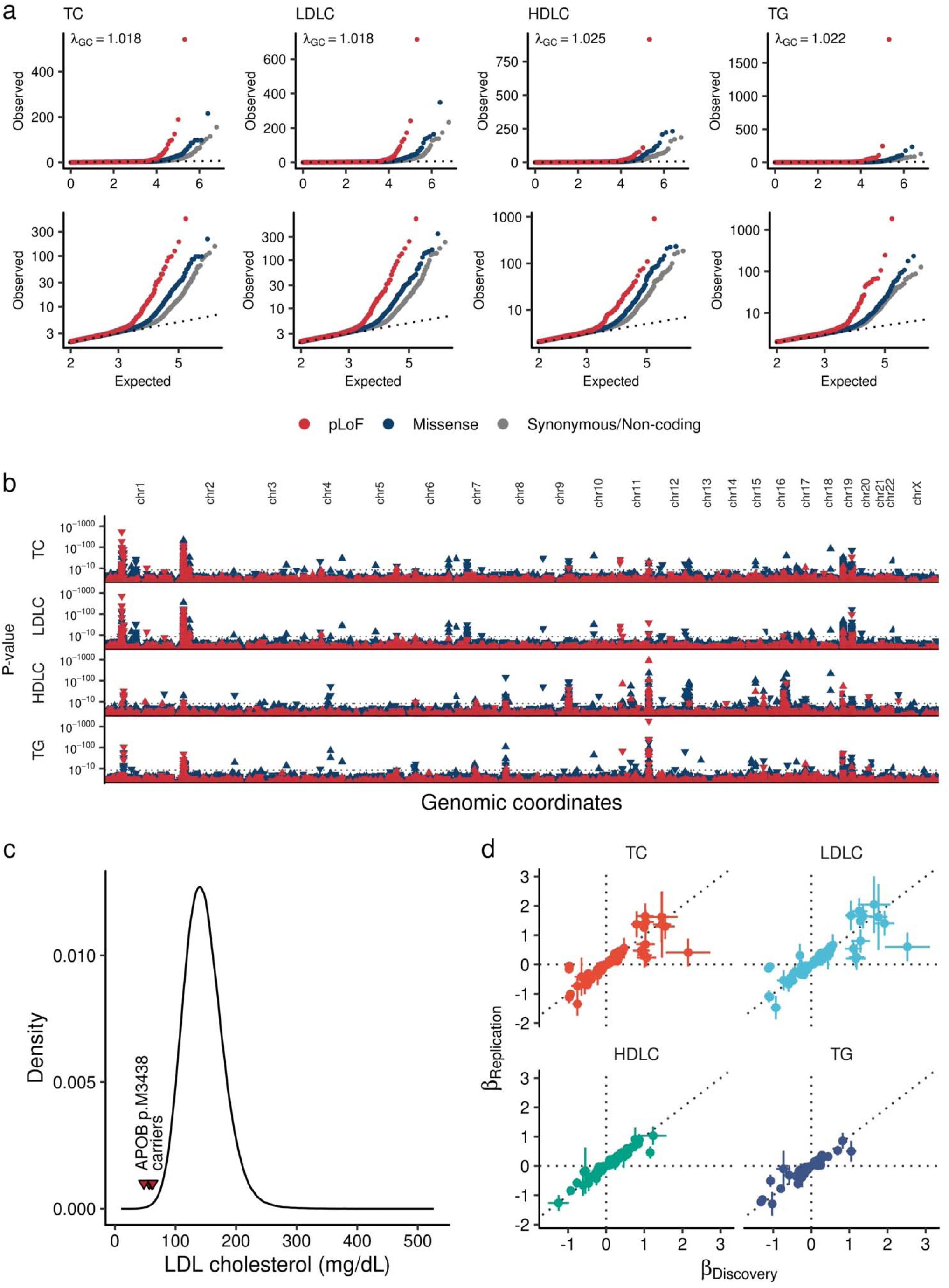
Exome wide association analysis over a million individuals. **a.** Quantile-quantile plot. Upper panels are quantile-quantile plots for four tested lipid traits. Each dot indicates a tested variant. Colors indicate variant annotation. Dotted lines show expected distribution. Lower panel focused variants with expected *P*-value < 0.01. **b.** 184 exome wide significant loci. The horizontal axis shows genomic coordinates, and the vertical axis shows *P*-values. The red triangles indicate pLoF variant and blue indicate missense variant. The upward triangles indicate trait increasing associations, and downward triangles indicate trait decreasing associations. The *P*-values were calculated by linear mixed model with two-sided test. The *P*-values were not adjusted for multiple testing correction. **c.** Penetrant association of APOB p.M3438X. The curve indicates LDLC distribution of the European-like population in the UKB (N = 409,046). The red triangles indicate LDLC level of the carriers of APOB p.M3438X. **d.** Replication evidence in the independent study for associated variants. Each point represents rare-coding genetic variants that are significantly associated with blood lipids in this study. The horizontal axes display the effect sizes from this study (Discovery, N_MAX_ = 1,057,837), while the vertical axes present the effect sizes from the previous exome-array study (Replication, N_MAX_ = 358,251, Lu et al., *Nat Genet* 2017). The error bars represent the 95% confidence intervals in each study. TC, Total Cholesterol; LDLC, Low Density Lipoprotein Cholesterol; HDLC, High density lipoprotein cholesterol; TG, Triglycerides; GC; Genomic Control; pLoF, predicted Loss of Function; Chr, Chromosome.

**Extended Data Figure 3.**
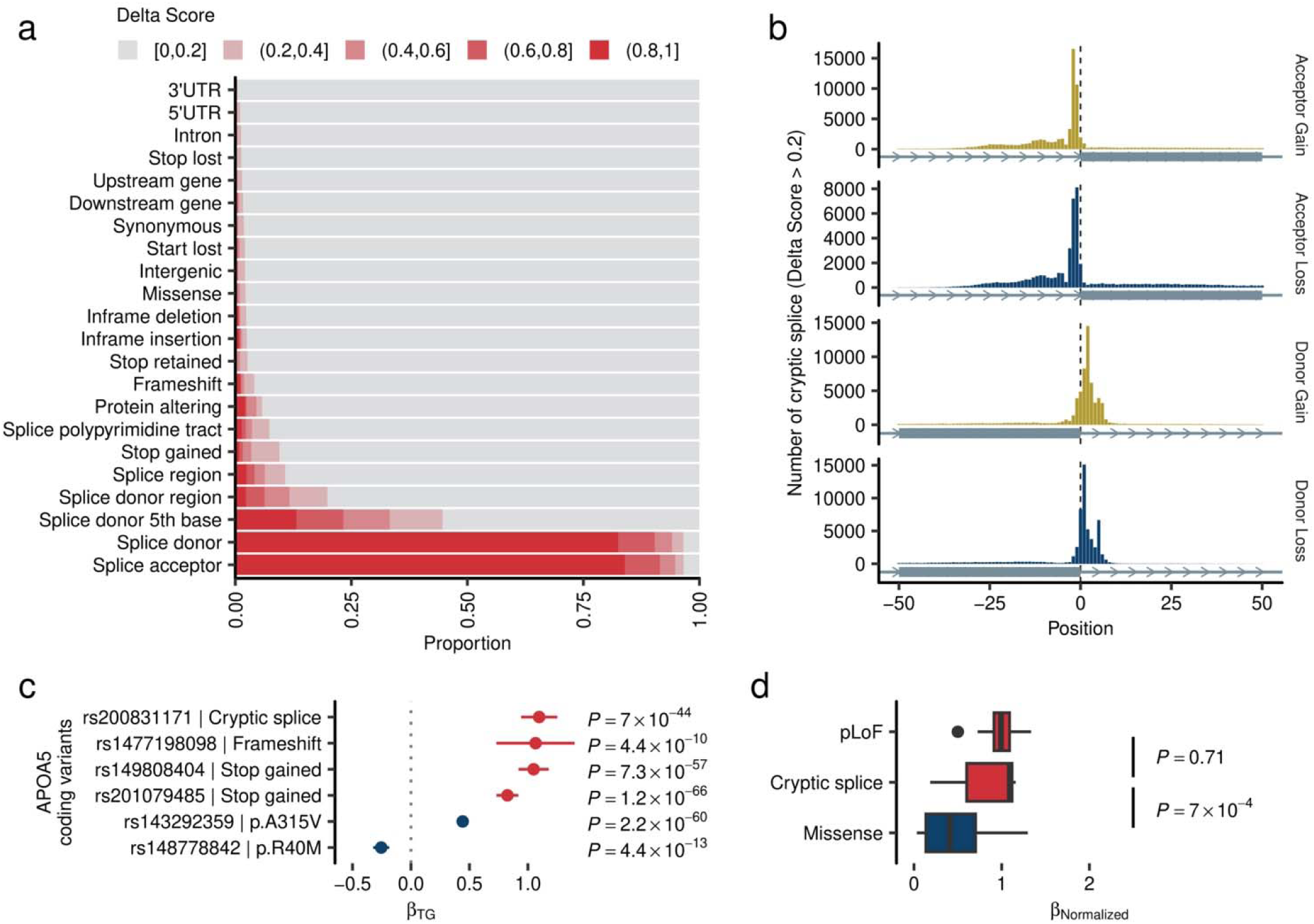
Cryptic splice variants affect human blood lipids. **a.** Distribution of cryptic splice variants across canonical variant classes. The bar graphs illustrate the proportion of cryptic splice variants within the canonical annotations, with the colors of the bars indicating the Delta Score (DS). **b.** Distribution of cryptic splice variants around exon-intron boundary. The histogram shows the positions of cryptic splicing variants (DS > 0.8) in relation to the exon-intron boundary. Exons are represented by blue rectangles. **c.** Strong expressivity of *APOA5* cryptic splice variant. Each dot indicates effect size of variant calculated by linear mixed model. The unit of effect size is a standard error of blood triglycerides. The error bar indicates 95% confidence interval of effect size. Red dots indicate pLoF variants and blue dots indicate missense variants. The *P*-values were calculated by linear mixed model with two-sided test. The *P*-values were not adjusted for multiple testing correction. **d.** Strong expressivity of Cryptic splice variants. The horizontal axis shows the normalized effect sizes for pLoF, pLoF (cryptic splice) and missense variants. The analysis was restricted to the genes both harboring pLoF, cryptic splice, and missense variants. Boxplot shows the median value as the centerline; box boundaries show the first and third quartiles and whiskers extending 1.5 times the interquartile range. The *P*-values were calculated by the Wilcoxon rank-sum test. The *P*-values were not adjusted for multiple testing correction. UTR, Untranslated Region; pLoF, predicted Loss of Function; TG, Triglycerides.

**Extended Data Figure 4.**
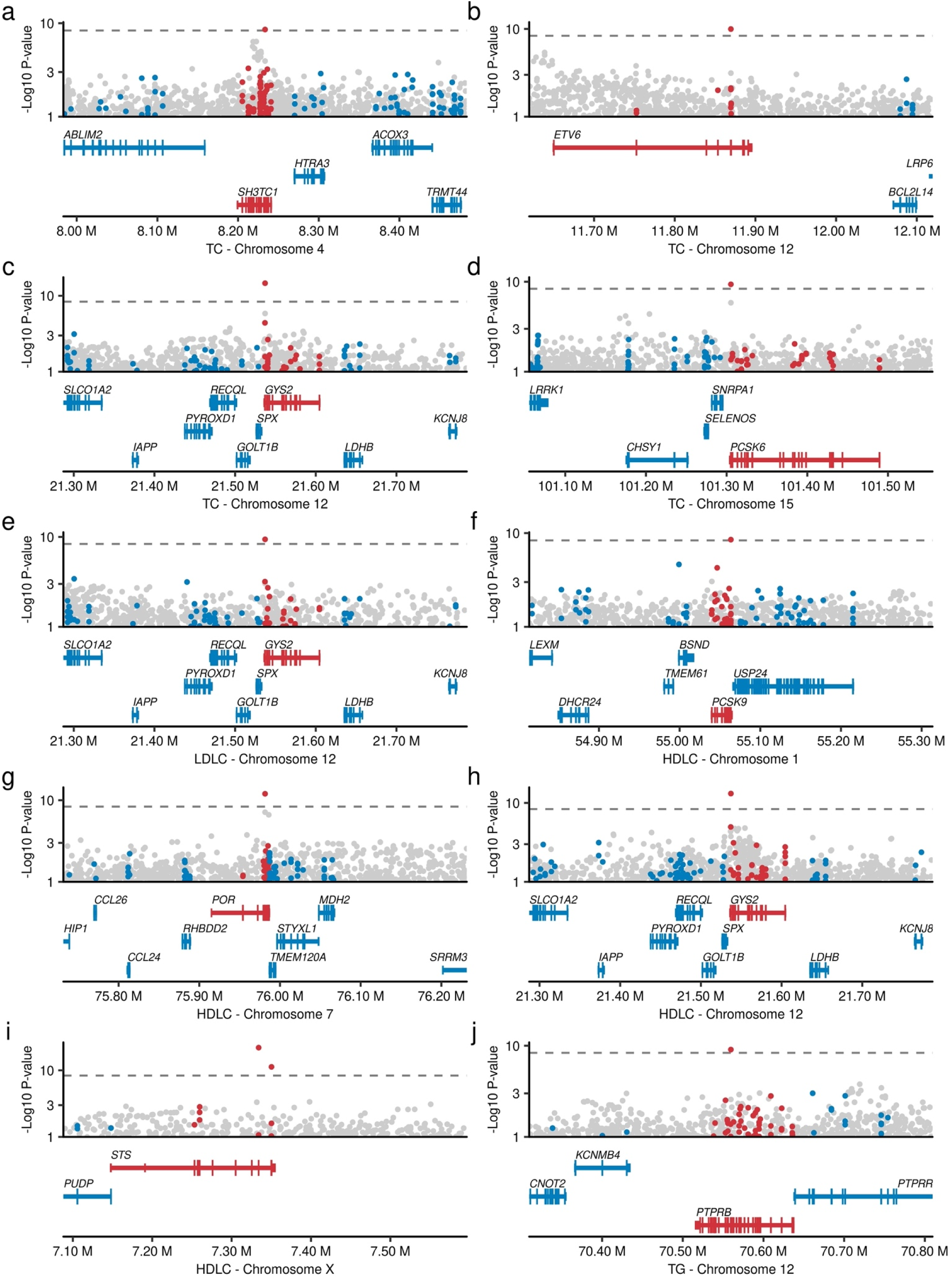
Novel loci driven by rare coding variants. The horizontal axes represent genomic coordinates, while the vertical axes denote the negative log10 P-values. Red dots illustrate the association of rare coding variants in genes with significant variants. In contrast, blue dots show the association of rare coding variants in genes without significant variants. Gray dots represent common variant associations from a previous study (Graham et al., *Nature* 2021). The dashed line in the upper panel indicates the exome-wide significance threshold (*P* < 4.4 × 10^−9^). The lower panel illustrates the coding genes within the locus; genes harboring significant variants are highlighted in red, and others are in blue. The *P*-values were calculated by linear mixed model with two-sided test. The *P*-values were not adjusted for multiple testing correction. TC, Total Cholesterol; LDLC, Low Density lipoprotein cholesterol; HDLC, High Density Lipoprotein Cholesterol; TG, Triglycerides.

**Extended Data Figure 5.**
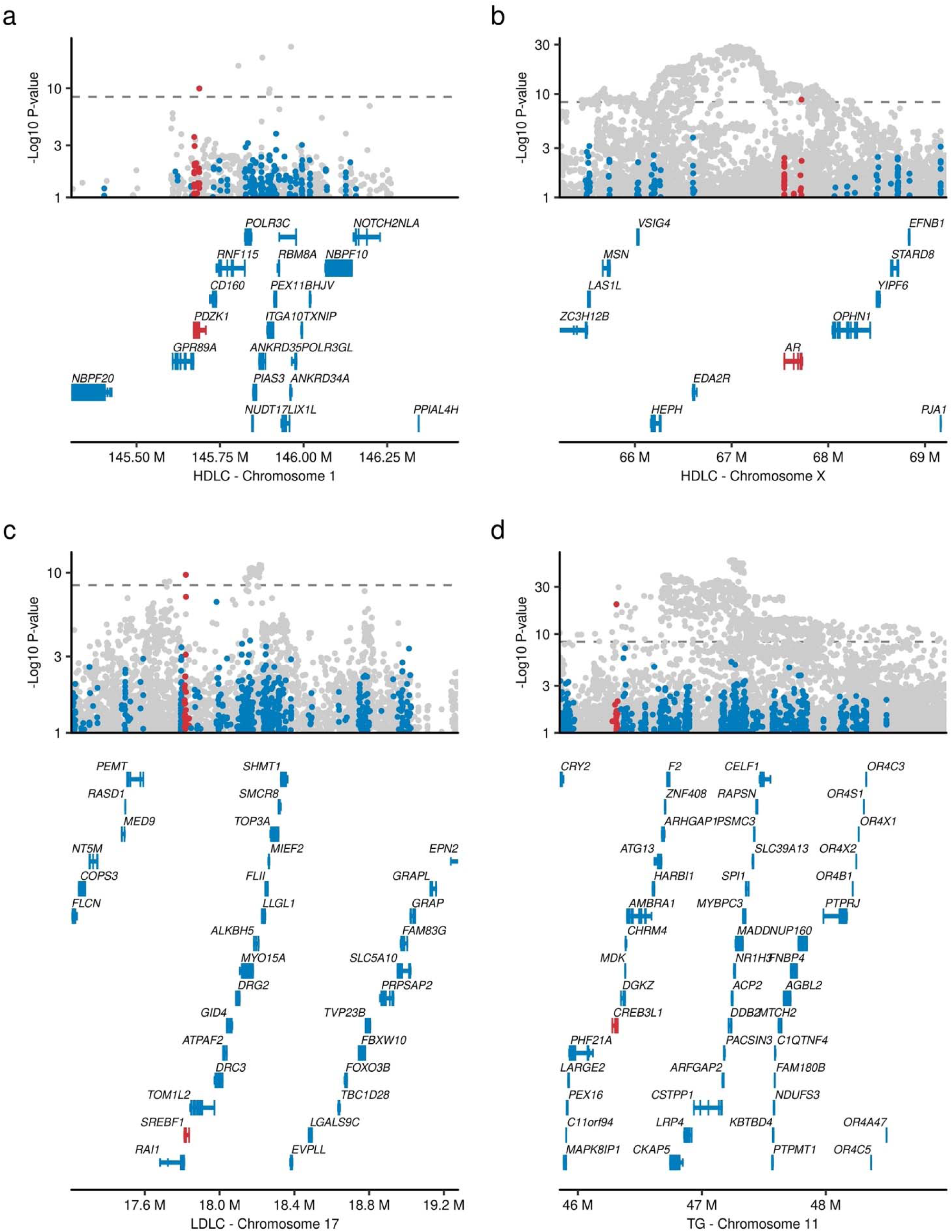
Implicated causal genes in the established lipid associated loci. The horizontal axes represent genomic coordinates, while the vertical axes denote the negative log10 P-values for *PDZK1* (**a**), *SREBF1* (**b**), *AR* (**c**), and *CREB3L1* (**d**). Red dots illustrate the association of rare coding variants in genes with significant variants. In contrast, blue dots show the association of rare coding variants in genes without significant variants. Gray dots represent common variant associations from a previous study (Graham et al., *Nature* 2021). The dashed line in the upper panel indicates the exome-wide significance threshold (*P* < 4.4 × 10^−9^). The lower panel illustrates the coding genes within the locus; genes harboring significant variants are highlighted in red, and others are in blue. The *P*-values were calculated by linear mixed model with two-sided test. The *P*-values were not adjusted for multiple testing correction. The LDLC, low-density lipoprotein cholesterol; HDLC, high-density lipoprotein cholesterol; MVP, Million Veteran Program; UKB, UK Biobank; AFR, African-like population; EUR, European-like population; HIS, Hispanic-like population.

**Extended Data Figure 6.**
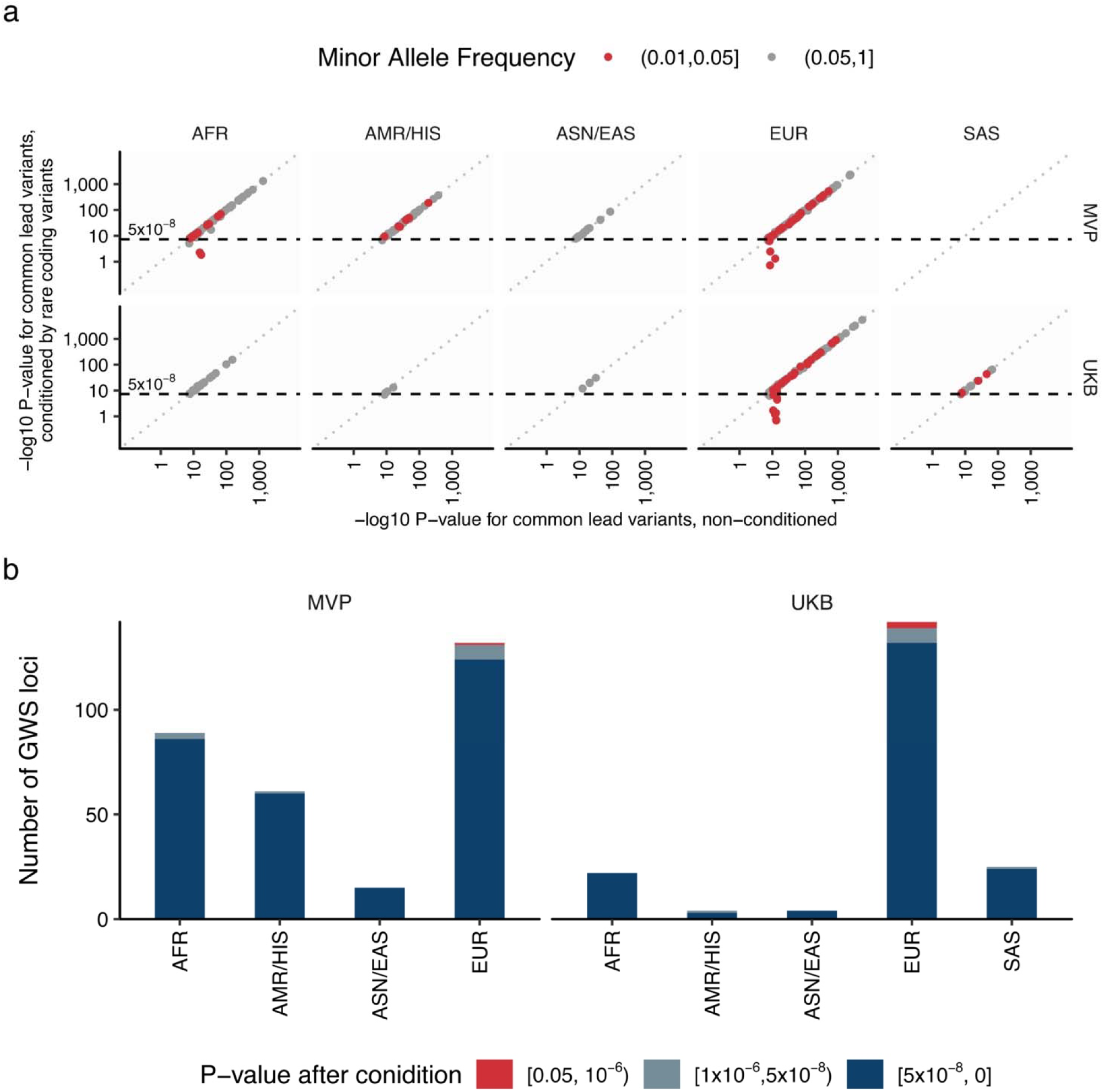
Independence of common genetic signals and rare genetic signals. **a.** Each dot indicates common genetic variant (MAF ≥ 1%) associated with blood lipids within the loci identified by rare genetic associations in this study. We compare non-conditioned and conditioned statistics in this figure to assess the independence of common genetic signals and rare genetic signals. In conditioned analysis, we introduced all the associated rare variant genotypes as covariates in the linear regression model (Methods and Supplementary Notes V). The horizontal axes show -log_10_ P-values without conditioning and the vertical axes show them with conditioning by rare variant genotypes with EWS. The *P*-values were calculated by linear regression model with two-sided test. The *P*-values were not adjusted for multiple testing correction. **b.** The number of common genetic signals affected by rare genetic signals were summarized in the bar chart. The bar chart indicates number of common genetic signals, and the color classifies the signals based on the P-values of common genetic signals after conditioning by rare genetic signals. MVP, Million Veteran Program; UKB, UK Biobank; AFR, African-like population; AMR, Admixed-American-like population; ASN, Asian-like population; EAS, East-Asian-like population; EUR, European-like population; HIS, Hispanic-like population; SAS, South-Asian-like population; TC, Total Cholesterol; LDLC, Low Density Lipoprotein Cholesterol; HDLC, High Density Lipoprotein Cholesterol; TG, Triglycerides.

**Extended Data Figure 7.**
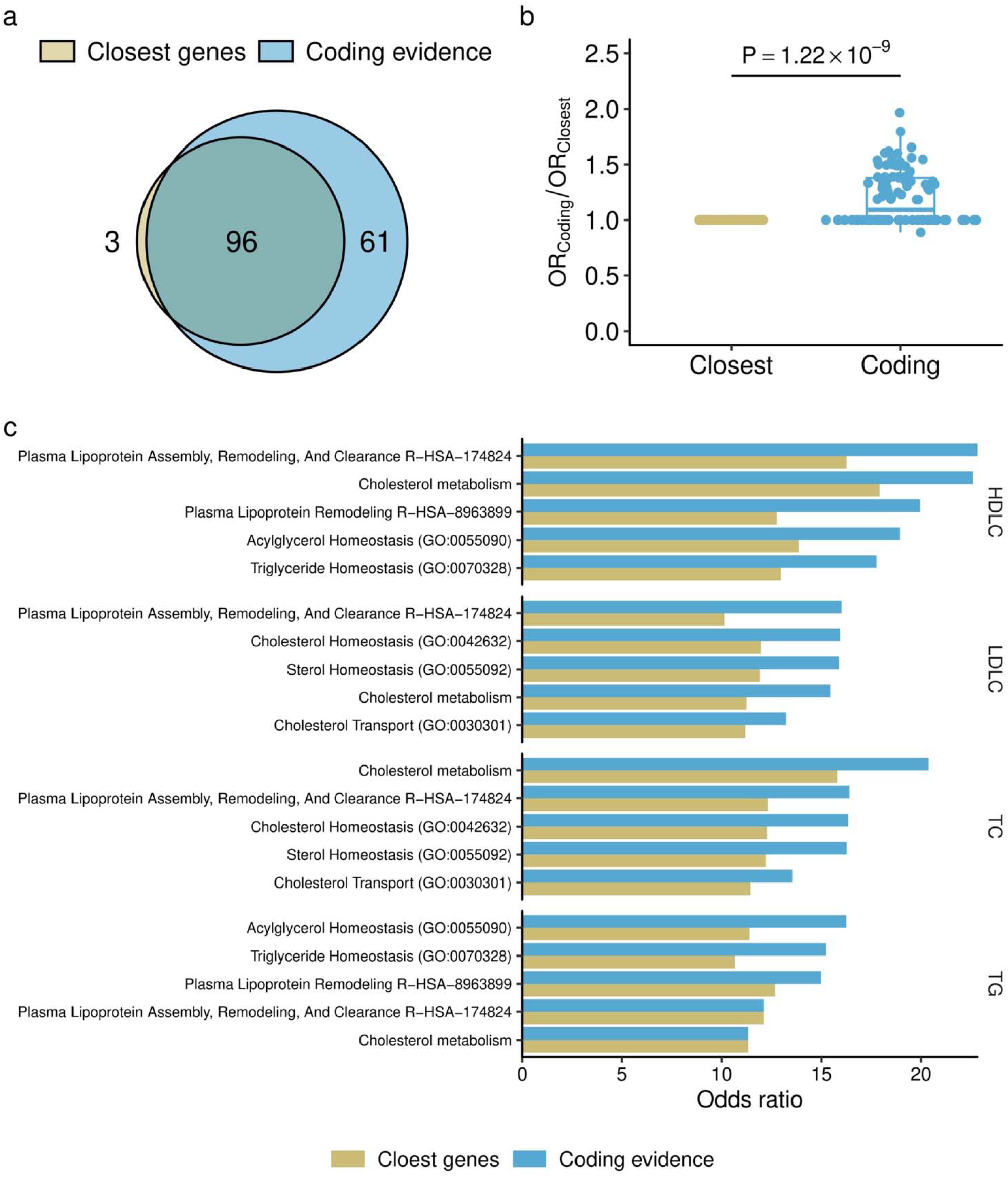
Enhanced enrichment of associated genes in the causal pathway. **a.** Pathway enrichment by common and rare genetic signals. Venn diagram showing significantly enriched pathways for gene sets based on common and rare variant associations. The gene set for common variants was defined by the nearest genes to the lead common variant (Graham et al., *Nature* 2021), while the gene set for rare variants was defined by the genes harboring exome-wide significant (EWS) associations in this study. **b.** Pairwise comparison of odds ratios for gene sets (n = 96) associated with both common and rare variants. The vertical axis shows the relative odds ratio (OR_Common_/OR_Rare_). The *P*-value was computed by paired Wilcoxson’s rank-sum test. Boxplot shows the median value as the centerline; box boundaries show the first and third quartiles and whiskers extending 1.5 times the interquartile range. **c.** Pathway enrichment analysis was performed on genes harboring rare coding variants associated with lipids and on genes closest to common variant associations with blood lipids. The top five enriched pathways for each trait are displayed. The horizontal axis denotes the odds ratio, with red bars indicating the odds ratios for the gene set with rare variants and blue bars for the gene set with common variants. GO, Gene Ontology; TC, Total Cholesterol; HDLC, High Density Lipoprotein Cholesterol; LDLC, Low Density Lipoprotein Cholesterol; TG, Triglycerides.

**Extended Data Figure 8.**
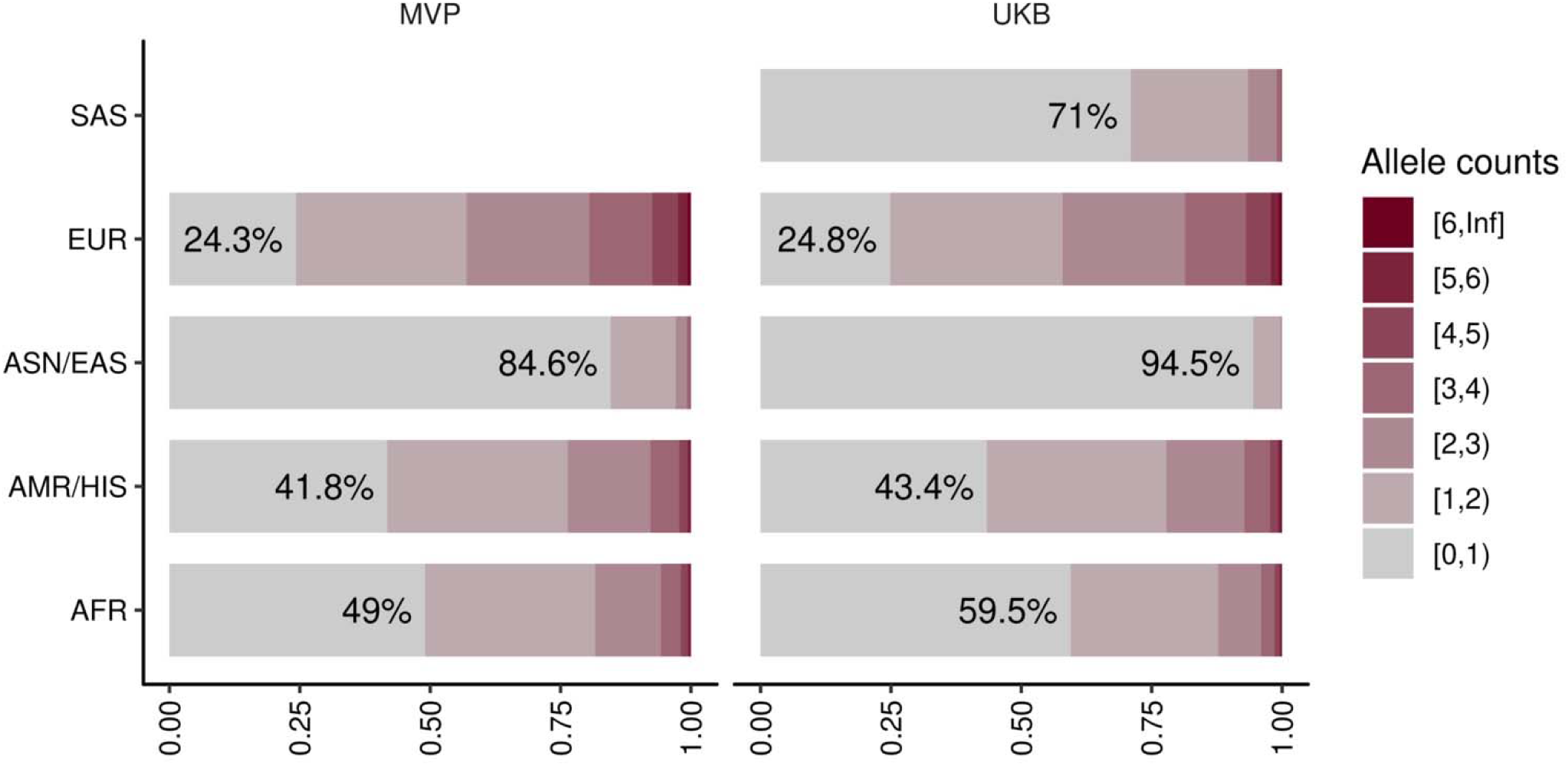
Limited discovery in non-European lipid associated alleles. This figure shows the proportion of the individuals with lipid associated alleles identified in this study. The colors of bar charts indicate allele counts of lipid associated alleles possessed by individuals. The percentages in the bars are showing the proportion of the individuals without lipid associated alleles in the population. MVP, Million Veteran Program; UKB, UK-Biobank; AFR, African-like population, AMR, Admixed-American-like population, HIS, Hispanic-like population, ASN, Asian-like population; EAS, East-Asian-like population; EUR, European-like population; SAS, South-Asian-like population.

**Extended Data Figure 9.**
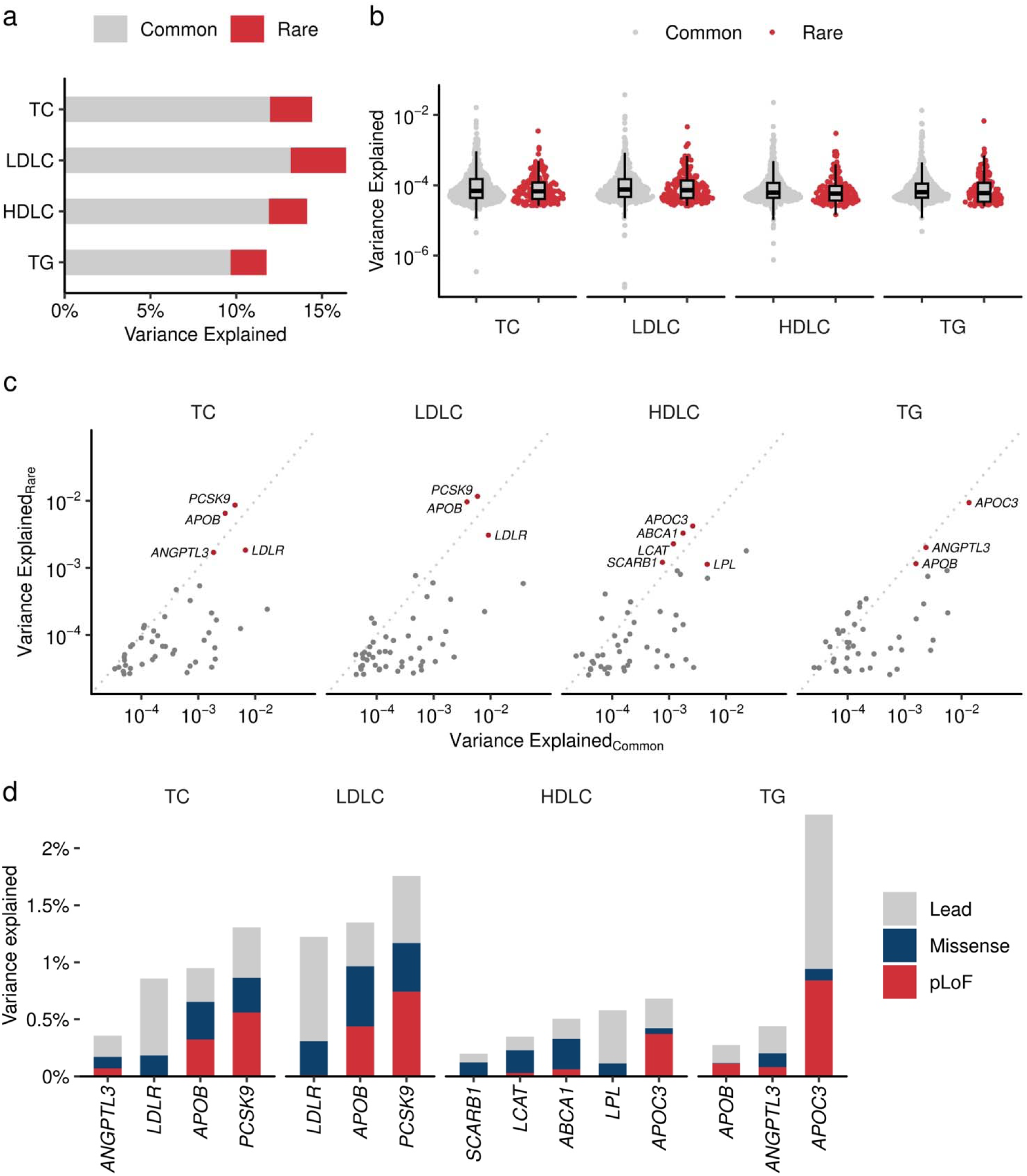
Contribution of rare coding variants to trait variance. **a.** Phenotypic variance explained (PVE) by common and rare variants. The height of the bar chart indicates the PVE by GWAS lead variant (yellow) and the sum of rare coding variants in the locus (dark blue). PVE is computed by the formula *2f(1-f)*β^2^, where *f* is the allele frequency and β is the effect size. **b.** PVE by individual variants. Grey dots indicate common (Grahan et al. Nature 2021) and red dots indicate rare (current study) variants. Boxplot shows the median value as the centerline; box boundaries show the first and third quartiles and whiskers extending 1.5 times the interquartile range. **c.** Trait variance by rare coding variant and common genetic signals. The horizontal axis indicates PVE by lead variant in the GWAS loci. The vertical axis indicates the sum of PVEs by rare coding variants in the locus. **d.** The cumulative contribution of lead and rare coding variants for trait variance. PVE by each rare variant in representative genes. Lead variant in the locus in gray, the sum of PVEs by pLoF in red and missense in dark blue. PVE, Phenotypic Variance Explained; GWAS, Genome Wide Association Study. TC, Total Cholesterol; High Density Lipoprotein Cholesterol; LDLC, Low Density Lipoprotein Cholesterol; TG, Triglycerides; pLoF, predicted Loss of Function.

## References

1. Wiegman, A., et al. Familial hypercholesterolaemia in children and adolescents: gaining decades of life by optimizing detection and treatment. European Heart Journal 36, 2425–2437 (2015).

2. Gidding, S.S., et al. The Agenda for Familial Hypercholesterolemia: A Scientific Statement From the American Heart Association. Circulation 132, 2167–2192 (2015).

3. Versmissen, J., et al. Efficacy of statins in familial hypercholesterolaemia: a long term cohort study. BMJ 337, a2423 (2008).

4. Neil, A., et al. Reductions in all-cause, cancer, and coronary mortality in statin-treated patients with heterozygous familial hypercholesterolaemia: a prospective registry study. European Heart Journal 29, 2625–2633 (2008).

5. Pijlman, A.H., et al. Evaluation of cholesterol lowering treatment of patients with familial hypercholesterolemia: a large cross-sectional study in The Netherlands. Atherosclerosis 209, 189–194 (2010).

6. Nordestgaard, B.G., et al. Familial hypercholesterolaemia is underdiagnosed and undertreated in the general population: guidance for clinicians to prevent coronary heart disease: Consensus Statement of the European Atherosclerosis Society. European Heart Journal 34, 3478–3490 (2013).

7. Sturm, A.C., et al. Clinical Genetic Testing for Familial Hypercholesterolemia: JACC Scientific Expert Panel. J Am Coll Cardiol 72, 662–680 (2018).

8. Benn, M., Watts, G.F., Tybjaerg-Hansen, A. & Nordestgaard, B.G. Mutations causative of familial hypercholesterolaemia: screening of 98 098 individuals from the Copenhagen General Population Study estimated a prevalence of 1 in 217. Eur Heart J 37, 1384–1394 (2016).

9. Natarajan, P., et al. Aggregate penetrance of genomic variants for actionable disorders in European and African Americans. Science Translational Medicine 8, 364ra151–364ra361 (2016).

10. Sun, Y.V., et al. Effects of Genetic Variants Associated with Familial Hypercholesterolemia on Low-Density Lipoprotein-Cholesterol Levels and Cardiovascular Outcomes in the Million Veteran Program. Circulation: Genomic and Precision Medicine 11(2018).

11. Forrest, I.S., et al. Population-Based Penetrance of Deleterious Clinical Variants. JAMA 327, 350 (2022).

12. Clarke, S.L., et al. Coronary Artery Disease Risk of Familial Hypercholesterolemia Genetic Variants Independent of Clinically Observed Longitudinal Cholesterol Exposure. Circ Genom Precis Med 15, e003501 (2022).

13. Dewey, F.E., et al. Distribution and clinical impact of functional variants in 50,726 whole-exome sequences from the DiscovEHR study. Science 354, aaf6814 (2016).

14. Green, R.C., et al. ACMG recommendations for reporting of incidental findings in clinical exome and genome sequencing. Genetics in Medicine 15, 565–574 (2013).

15. Richards, S., et al. Standards and guidelines for the interpretation of sequence variants: a joint consensus recommendation of the American College of Medical Genetics and Genomics and the Association for Molecular Pathology. Genetics in Medicine 17, 405–424 (2015).

16. Blout Zawatsky, C.L., et al. Returning actionable genomic results in a research biobank: Analytic validity, clinical implementation, and resource utilization. The American Journal of Human Genetics 108, 2224–2237 (2021).

17. Sharo, A.G., Zou, Y., Adhikari, A.N. & Brenner, S.E. ClinVar and HGMD genomic variant classification accuracy has improved over time, as measured by implied disease burden. Genome Medicine 15(2023).

18. Kessler, M.D., et al. Challenges and disparities in the application of personalized genomic medicine to populations with African ancestry. Nature Communications 7, 12521 (2016).

19. Manrai, A.K., et al. Genetic Misdiagnoses and the Potential for Health Disparities. New England Journal of Medicine 375, 655–665 (2016).

20. Hunter-Zinck, H., et al. Genotyping Array Design and Data Quality Control in the Million Veteran Program. Am J Hum Genet 106, 535–548 (2020).

21. Bycroft, C., et al. The UK Biobank resource with deep phenotyping and genomic data. Nature 562, 203–209 (2018).

22. Taliun, D., et al. Sequencing of 53,831 diverse genomes from the NHLBI TOPMed Program. Nature 590, 290–299 (2021).

23. Backman, J.D., et al. Exome sequencing and analysis of 454,787 UK Biobank participants. Nature 599, 628–634 (2021).

24. Bick, A.G., et al. Genomic data in the All of Us Research Program. Nature 627, 340–346 (2024).

25. Jaganathan, K., et al. Predicting Splicing from Primary Sequence with Deep Learning. Cell 176, 535–548 e524 (2019).

26. Lu, X., et al. Exome chip meta-analysis identifies novel loci and East Asian– specific coding variants that contribute to lipid levels and coronary artery disease. Nature Genetics 49, 1722–1730 (2017).

27. Karczewski, K.J., et al. The mutational constraint spectrum quantified from variation in 141,456 humans. Nature 581, 434–443 (2020).

28. McLaren, W., et al. The Ensembl Variant Effect Predictor. Genome Biol 17, 122 (2016).

29. Liu, X., Li, C., Mou, C., Dong, Y. & Tu, Y. dbNSFP v4: a comprehensive database of transcript-specific functional predictions and annotations for human nonsynonymous and splice-site SNVs. Genome Med 12, 103 (2020).

30. Allard, D., et al. Novel mutations of thePCSK9 gene cause variable phenotype of autosomal dominant hypercholesterolemia. Human Mutation 26, 497–497 (2005).

31. Hopkins, P.N., et al. Characterization of Autosomal Dominant Hypercholesterolemia Caused by *PCSK9* Gain of Function Mutations and Its Specific Treatment With Alirocumab, a PCSK9 Monoclonal Antibody. Circulation: Cardiovascular Genetics 8, 823–831 (2015).

32. Aguet, F., et al. The GTEx Consortium atlas of genetic regulatory effects across human tissues. Science 369, 1318–1330 (2020).

33. Orho, M., et al. Mutations in the liver glycogen synthase gene in children with hypoglycemia due to glycogen storage disease type 0. Journal of Clinical Investigation 102, 507–515 (1998).

34. Reed, M.J., Purohit, A., Woo, L.W.L., Newman, S.P. & Potter, B.V.L. Steroid Sulfatase: Molecular Biology, Regulation, and Inhibition. Endocrine Reviews 26, 171–202 (2005).

35. Assanasen, C., et al. Cholesterol binding, efflux, and a PDZ-interacting domain of scavenger receptor–BI mediate HDL-initiated signaling. Journal of Clinical Investigation 115, 969–977 (2005).

36. Goldstein, J.L., Debose-Boyd, R.A. & Brown, M.S. Protein Sensors for Membrane Sterols. Cell 124, 35–46 (2006).

37. Ehrhardt, N., et al. Hepatic Tm6sf2 overexpression affects cellular ApoB-trafficking, plasma lipid levels, hepatic steatosis and atherosclerosis. Human Molecular Genetics 26, 2719–2731 (2017).

38. Pauling, L., Itano, H.A., Singer, S.J. & Wells, I.C. Sickle Cell Anemia, a Molecular Disease. Science 110, 543–548 (1949).

39. Ingram, V.M. A Specific Chemical Difference Between the Globins of Normal Human and Sickle-Cell Anæmia Hæmoglobin. Nature 178, 792–794 (1956).

40. Xia, W., et al. Loss of ABHD15 Impairs the Anti-lipolytic Action of Insulin by Altering PDE3B Stability and Contributes to Insulin Resistance. Cell Reports 23, 1948–1961 (2018).

41. Landrum, M.J., et al. ClinVar: improving access to variant interpretations and supporting evidence. Nucleic Acids Res 46, D1062–D1067 (2018).

42. Zou, H., Yang, N., Zhang, X. & Chen, H.-W. RORγ is a context-specific master regulator of cholesterol biosynthesis and an emerging therapeutic target in cancer and autoimmune diseases. Biochemical Pharmacology 196, 114725 (2022).

43. Cai, D., et al. RORγ is a targetable master regulator of cholesterol biosynthesis in a cancer subtype. Nature Communications 10(2019).

44. Jurgens, S.J., et al. Rare coding variant analysis for human diseases across biobanks and ancestries. Nature Genetics 56, 1811–1820 (2024).

45. Barton, A.R., Sherman, M.A., Mukamel, R.E. & Loh, P.-R. Whole-exome imputation within UK Biobank powers rare coding variant association and fine-mapping analyses. Nature Genetics 53, 1260–1269 (2021).

46. Heyne, H.O., et al. Mono- and biallelic variant effects on disease at biobank scale. Nature 613, 519–525 (2023).

47. Halford, J.L., et al. Endophenotype effect sizes support variant pathogenicity in monogenic disease susceptibility genes. Nature Communications 13(2022).

48. Sun, K.Y., et al. A deep catalog of protein-coding variation in 985,830 individuals. bioRxiv, 2023.2005.2009.539329 (2023).

49. Koyama, S., et al. Decoding Genetics, Ancestry, and Geospatial Context for Precision Health. medRxiv, 10.1101/2023.1110.1124.23297096 (2023).

50. Graham, S.E., et al. The power of genetic diversity in genome-wide association studies of lipids. Nature 600, 675–679 (2021).

51. Verma, A., et al. Diversity and Scale: Genetic Architecture of 2,068 Traits in the VA Million Veteran Program. (Cold Spring Harbor Laboratory, 2023).

52. Klarin, D., et al. Genetics of blood lipids among ∼300,000 multi-ethnic participants of the Million Veteran Program. Nature genetics 50, 1514–1523 (2018).

53. Clarke, S.L., et al. Race and Ethnicity Stratification for Polygenic Risk Score Analyses May Mask Disparities in Hispanics. Circulation 146, 265–267 (2022).

54. Chang, C.C., et al. Second-generation PLINK: rising to the challenge of larger and richer datasets. Gigascience 4, 7 (2015).

55. Poterba, T., et al. The Scalable Variant Call Representation: Enabling Genetic Analysis Beyond One Million Genomes. bioRxiv, 10.1101/2024.1101.1109.574205 (2024).

56. Li, H. Toward better understanding of artifacts in variant calling from high-coverage samples. Bioinformatics 30, 2843–2851 (2014).

57. Poplin, R., et al. A universal SNP and small-indel variant caller using deep neural networks. Nature Biotechnology 36, 983–987 (2018).

58. Fang, H., et al. Harmonizing Genetic Ancestry and Self-identified Race/Ethnicity in Genome-wide Association Studies. The American Journal of Human Genetics 105, 763–772 (2019).

59. Ross, P.B., Song, J., Tsao, P.S. & Pan, C. Trellis for efficient data and task management in the VA Million Veteran Program. Scientific Reports 11(2021).

60. Mbatchou, J., et al. Computationally efficient whole-genome regression for quantitative and binary traits. Nat Genet 53, 1097–1103 (2021).

61. Mägi, R. & Morris, A.P. GWAMA: software for genome-wide association meta-analysis. BMC Bioinformatics 11, 288 (2010).

62. Frankish, A., et al. GENCODE reference annotation for the human and mouse genomes. Nucleic Acids Res 47, D766–D773 (2019).

63. Xie, Z., et al. Gene Set Knowledge Discovery with Enrichr. Curr Protoc 1, e90 (2021).

64. Liu, D.J., et al. Exome-wide association study of plasma lipids in >300,000 individuals. Nature Genetics 49, 1758–1766 (2017).

65. McKenna, A., et al. The Genome Analysis Toolkit: a MapReduce framework for analyzing next-generation DNA sequencing data. Genome research 20, 1297–1303 (2010).

