## Supplementary Information I-VI for "Exome wide association study for blood lipids in 1,158,017 individuals from diverse populations"

Satoshi Koyama et al.

1. Supplementary Notes	2
   I. Imputation accuracy of rare variants in MVP
   II. Variant coverage
   III. Cryptic splice annotation
   IV. Power calculation
   V. Independent association of rare coding variants from common variants for lipids
   VI. Enhanced gene set enrichment by rare coding associations for lipids
2. Supplementary References	13
   **Supplementary Notes**

**I. Imputation accuracy of rare variants in MVP**

We imputed MVP cohort up to TOPMed imputation reference panel (r2) including 97,256 individuals’ genome and 308,107,085 variants from diverse populations. To assess the imputation accuracy in MVP, we compare the imputed dosage and sequenced genotypes by high depth whole genome sequencing data. We selected 3,594 African-like population and 6,342 European-like population and compared imputed dosages and genotypes determined by deep coverage WGS. We restricted analysis to the high-quality variants in WGS by focusing on i) SNVs, ii) High genotyping rate (missingness < 1%), iii) Hardy Weinberg equilibrium P-value < 1 × 10^−6^, iv) not in Low-complexity regions, resulting in 6,767,236 SNVs for the African American-like population dataset and 7,278,589 SNVs for the European-like population dataset. From the imputed dataset, we extracted individual-level imputed dosages for the same variants found in the WGS dataset and assessed genotypic concordance. If the variants were not found in the imputed dataset, these variants were all set as reference homozygotes. In addition to traditional measurements for imputation quality (R^2^, Pearson’s correlation coefficient between imputed dosage and sequenced genotypes), we computed rare-variant focused metrics by converting imputed dosage (continuous value [0 - 2]) and sequenced genotypes (0, 1, 2) into dichotomized genotypes (0 = [0 - 0.5), 1 = [0.5 - 2] for imputed dosage and 0 = 0, 1 = 1 or 2 for sequenced genotypes). Using such dichotomized genotypes, we computed i) True Positive Rate [TPR, True positive/(True positive + False Negative)], and ii) False Positive Rate [FPR, False Positive/(False Positive + True Negative)] for each variant. We also performed the analysis for the down-sampled European dataset (N = 3,594). We summarized distributions of R^2^, TPR, FPR by computing mean by MAC/MAF bins (Extended Data Figure 1a). We observed a high level of genotypic concordance between imputed and sequenced genotypes for the variants with MAC ≥ 5 (mean TPR > 82% and mean FPR < 5.3%) in both African-like and European-like datasets

**II. Variant coverage**

To compute possible protein-coding variants, canonical transcript for 19,603 protein coding transcripts defined in Gencode v41 were used. We identified 200,981 exons spanning 71,401,590 sites. For each site identified, we simulated all possible single nucleotide substitution (e.g. For a site with reference allele of A, we generated 3 different variants defined by alternate allele as C, G, T) on exon and +/− 2 base pairs and annotated by VEP version 107. Among 214,204,770 variants we simulated, we identified 6,654,122 possible pLoF SNVs and 73,074,537 missense SNVs. Among these all possible SNVs in our coding gene model, we computed the coverage by dataset we used. Overall, UKB whole exome sequencing (WES) data from EUR showed better coverage (1.04% of all pLoF SNVs and 2.32% of all missense SNVs) than MVP imputed data from EUR (0.68% and 1.55%). However, diverse populations included in the MVP substantially expand allelic diversity and showed equivalent coverage when we considered all populations (1.10% in UKB and 0.98% in MVP for pLoF, and 2.54% in UKB and 2.30% in MVP for missense). Combining MVP, UKB and AOU, the identified variants included 115,284 pLoF SNVs (1.73%) and 2,784,672missense SNVs (3.81%) among these possible coding variants (Supplementary Table 3). Please note that this analysis is restricted to SNVs and does not include insertions/deletions, which are difficult to simulate in all possible combinations.

**III. Cryptic splice annotation**

We employed Splice AI algorithms (https://github.com/Illumina/SpliceAI, Sept 6, 2021)^1^ to predict cryptic splice sites. In this study, we assessed various genetic variants and estimated their Delta Score (DS), which serves as a measure of their potential to induce cryptic splicing events—those that introduce non-canonical splicing patterns. Our analysis primarily focused on the coding transcript set (n = 19,603) we defined and used throughout in this study. We also utilized the "mask" feature to filter out scores that could either enhance splicing at canonical sites or reduce splicing at non-canonical sites. Following default recommendation, we set maximum distance between variant and cryptic splice site as 50 base pairs.

In total, we computed predictions for 9,645,514 variants found in the target region ± 50bp defined by UKB WES and successfully obtaining predictions for 96.9% (9,759,579/10,069,597) of variants. Splice AI provides four DS, ranging from 0 to 1, specifically for Donor Gain (DG), Donor Loss (DL), Acceptor Gain (AG), and Acceptor Loss (AL), along with their corresponding cryptic splice sites positions. To present a consolidated cryptic splice site annotation for each variant, we selected the maximum splice score (DS_Max_) for each variant.

We employed the cutoff for cryptic splice as Delta Score > 0.8 as a stringent cutoff suggested by the developer. The distribution of DS_Max_ scores is highly skewed, with approximately 73.7% (7,196,539/9,759,579) of variants predicted to have a DS_Max_ score of 0. Only a minority, comprising 0.598% (58,402/9,759,579) of the variants, exhibited DS_Max_ scores greater than 0.8.

83.2% of canonical splice site variants (Splice donor/acceptor) showed DS_Max_ > 0.8 and 96.6% showed DS_Max_ > 0.2. We identified enrichment of these variants in Splice region (intronic nucleotide 3 - 6 base away from exon-intron boundary, Extended Data Figure 3a) which were usually not considered as high impact on protein function. Particularly, in the 5^th^ base from splice donor (Splice donor 5^th^ base, SO:0001787) showed strong enrichment in variants with DS_Max_ > 0.8. Reflecting this functional potential, the variants in Splice donor 5^th^ base showed lower minor allele frequency than missense variants (Extended Data Figure 3b).

To maximize specificity, we decided to re-classify the variants with DS_Max_ score > 0.8 as pLoF variants and generate a new variant consequence – Cryptic splice in this study. By variant re-classification, the median MAF of Splice donor 5^th^ base was increased from 4.49 × 10^−5^ to 4.64 × 10^−5^, however, still higher than missense variants (4.69 × 10^−5^). The median MAF of cryptic splice variants was 4.00 × 10^−5^ and comparable with canonical pLoF variants.

**IV. Power calculation**

The power at specific allele frequency (*f* in the code), effect size (*beta* in the code), and significant threshold (*thr*) was computed by the following R code.

sigma	= sqrt(1 - 2*f*(1−f)*beta^2)

se	= sigma/sqrt(N*2*f*(1−f))

q.thr	= qchisq(thr, df = 1, ncp = 0, lower = F)

power	= pchisq(q.thr, df = 1, ncp = (beta/se)^2, lower = F)

(f, effect allele frequency; N, number of individuals; beta, effect sizes)

For variant-level power calculation, we estimated the empirical power for each variant based on its allele frequency and sample sizes in arbitrary effect sizes. We observed increase in sensitivity by increasing sample size from 100,000, to 500,000 and 1,000,000. For example, minimal allele frequency attained 80% of power to detect beta of at least 3 SD was decreased from 2.5 × 10^−5^ to 5.1 × 10^−6^, and 2.5 × 10^−6^. Also, detectable beta at MAF 0.01% was decreased from 1.5 to 0.672 and 0.476 (Extended Data Figure 1f).

For gene-level power calculation, we took the most powered variant for each gene and used it to represent the power of a gene. If we assume effect size of 3.0 SD per allele (i.e., largest effect sizes detected in this study), our study reached 80% power for 88.45% genes for pLoF associations and 95.3% of genes for missense associations. If we assume effect sizes of 1.0 SD per allele, 63.59% of genes for pLoF associations and 94.70% of genes for missense associations were sufficiently powered (Extended Data Figure 1g, Supplementary Table 5).

**V. Independent association of rare coding variants from common variants for lipids**

We conducted conditioning association analysis to test the independency between common genetic associations and rare genetic associations. First, we determined common variant association (MAF ≥ 1%) using TOPMed imputed genotype dosage in both MVP and UKB (non-conditioned analysis). Among 184 loci identified by rare coding variants, 143 and 142 loci archived genome wide significance (*P*_Unconditioned_ < 5 × 10^−8^) by common variants in MVP and UKB respectively. In the conditioned analysis, we introduced the genotypes of rare-coding variants with EWS identified in this study into the mixed model framework used in main analysis in this study. By conditioning rare coding signals, while 1 in MVP- and 3 in UKB- less common (MAF < 3%) lead variants substantially decreased its significance by rare variant conditioning (*P*_Conditioned_ ≥ 0.05), in the most of loci, the common variant signals were independent from the rare variant signals identified in this study (Extended Data Figures 6a and 6b). Overall, 95.1% (136/143) and 93.7% (133/143) loci did not lose genome wide significance by rare-variant conditioning (*P*_Conditioned_ < 5 × 10^−8^) in MVP and UKB respectively, suggesting independent causal mechanisms on the blood lipids.

**VI. Enhanced gene set enrichment by rare coding associations for lipids**

Compared to 99 pathways enriched by the nearest genes to the lead variants identified by recent GWAS, we identified 157 pathways enriched by genes with EWS with larger enrichment OR (*P* = 1.22 × 10^−9^, paired Wilcoxon Rank Sum test, Figure S6B). The associated genes were significantly enriched in cholesterol metabolism or related ontologies (Extended Data Figure 8c, Supplementary Table 12). These associated genes are also enriched under the regulation of several transcription factors previously implicated in lipid metabolism. One plausible example is the *CREB3L3* regulatory pathway with TG. Multiple *CREB3L3* pLoF/missense variants showed EWS with increased level of TG in this study (Supplementary Table 6) and others^2^. Consistent with these observations, we identified significant enrichment of the genes with EWS under the regulation of *CREB3L3* (*FGB, MLXIPL, HNF4A, APOE, ANGPTL4, APOB, PCK1, PPARA, A1CF, and TM6SF2*, OR = 24.3, *P* = 1.6 × 10^-10^). Other implicated transcription factors included LXR and RXR (TC, LDLC, TG), HNF4A (TC, LDLC,. TG), PPARA (LDLC, HDLC, TG), NR1I2 (Pregnane X receptor; PXR, TC, LDLC, HDLC, TG), NR1I3 (Constitutive androstane receptor; CAR, TC, LDLC, HDLC, TG), NR1H4 (Farnesoid X receptor; FXR, LDLC, HDLC, TG), NR0B2 (small heterodimer partner; SHP, LDLC, HDLC, TG).

**VA Million Veteran Program:
Core Acknowledgement for Publications
February 2024**

**MVP Program Office**

- Sumitra Muralidhar, Ph.D., Program Director
  US Department of Veterans Affairs, 810 Vermont Avenue NW, Washington, DC 20420
- Jennifer Moser, Ph.D., Associate Director, Scientific Programs
  US Department of Veterans Affairs, 810 Vermont Avenue NW, Washington, DC 20420
- Jennifer E. Deen, B.S., Associate Director, Cohort & Public Relations
  US Department of Veterans Affairs, 810 Vermont Avenue NW, Washington, DC 20420

**MVP Executive Committee**

- Co-Chair: Philip S. Tsao, Ph.D.
  VA Palo Alto Health Care System, 3801 Miranda Avenue, Palo Alto, CA 94304
- Co-Chair: Sumitra Muralidhar, Ph.D.
  US Department of Veterans Affairs, 810 Vermont Avenue NW, Washington, DC 20420
- J. Michael Gaziano, M.D., M.P.H.
  VA Boston Healthcare System, 150 S. Huntington Avenue, Boston, MA 02130
- Elizabeth Hauser, Ph.D.
  Durham VA Medical Center, 508 Fulton Street, Durham, NC 27705
- Amy Kilbourne, Ph.D., M.P.H.
  VA HSR&D, 2215 Fuller Road, Ann Arbor, MI 48105
- Michael Matheny, M.D., M.S., M.P.H.
  VA Tennessee Valley Healthcare System, 1310 24^th^ Ave. South, Nashville, TN 37212
- Dave Oslin, M.D.
  Philadelphia VA Medical Center, 3900 Woodland Avenue, Philadelphia, PA 19104

**MVP Co-Principal Investigators**

- J. Michael Gaziano, M.D., M.P.H.
  VA Boston Healthcare System, 150 S. Huntington Avenue, Boston, MA 02130
- Philip S. Tsao, Ph.D.
  VA Palo Alto Health Care System, 3801 Miranda Avenue, Palo Alto, CA 94304

**MVP Core Operations**

- Jessica V. Brewer, M.P.H., Director, MVP Recruitment & Enrollment
  VA Boston Healthcare System, 150 S. Huntington Avenue, Boston, MA 02130
- Mary T. Brophy M.D., M.P.H., Director, VA Central Biorepository
  VA Boston Healthcare System, 150 S. Huntington Avenue, Boston, MA 02130
- Kelly Cho, M.P.H, Ph.D., Director, MVP Phenomics Data Core
  VA Boston Healthcare System, 150 S. Huntington Avenue, Boston, MA 02130
- Lori Churby, B.S., Director, MVP Regulatory Affairs
  VA Palo Alto Health Care System, 3801 Miranda Avenue, Palo Alto, CA 94304
- Scott L. DuVall, Ph.D., Director, VA Informatics and Computing Infrastructure (VINCI)
  VA Salt Lake City Health Care System, 500 Foothill Drive, Salt Lake City, UT 84148
- Saiju Pyarajan Ph.D., Director, Data and Computational Sciences
  VA Boston Healthcare System, 150 S. Huntington Avenue, Boston, MA 02130
- Luis E. Selva, Ph.D., Executive Director, MVP Biorepositories
  VA Boston Healthcare System, 150 S. Huntington Avenue, Boston, MA 02130
- Shahpoor (Alex) Shayan, M.S., Director, MVP Recruitment and Enrollment Informatics
  VA Boston Healthcare System, 150 S. Huntington Avenue, Boston, MA 02130
- Stacey B. Whitbourne, Ph.D., Director, MVP Cohort Management
  VA Boston Healthcare System, 150 S. Huntington Avenue, Boston, MA 02130
- MVP Coordinating Centers
  - MVP Coordinating Center, Boston - J. Michael Gaziano, M.D., M.P.H.
    VA Boston Healthcare System, 150 S. Huntington Avenue, Boston, MA 02130
  - MVP Coordinating Center, Palo Alto – Philip S. Tsao, Ph.D.
    VA Palo Alto Health Care System, 3801 Miranda Avenue, Palo Alto, CA 94304
  - MVP Information Center, Canandaigua – Brady Stephens, M.S.
    Canandaigua VA Medical Center, 400 Fort Hill Avenue, Canandaigua, NY 14424
  - Cooperative Studies Program Clinical Research Pharmacy Coordinating Center, Albuquerque – Todd Connor, Pharm.D.; Dean P. Argyres, B.S., M.S.
    New Mexico VA Health Care System, 1501 San Pedro Drive SE, Albuquerque, NM 87108

**MVP Publications and Presentations Committee**

- Co-Chair: Themistocles L. Assimes, M.D., Ph. D
  VA Palo Alto Health Care System, 3801 Miranda Avenue, Palo Alto, CA 94304
- Co-Chair: Adriana Hung, M.D.; M.P.H
  VA Tennessee Valley Healthcare System, 1310 24^th^ Ave. South, Nashville, TN 37212
- Co-Chair: Henry Kranzler, M.D.
  Philadelphia VA Medical Center, 3900 Woodland Avenue, Philadelphia, PA 19104

**MVP Local Site Investigators**

- Samuel Aguayo, M.D., Phoenix VA Health Care System
  650 E. Indian School Road, Phoenix, AZ 85012
- Sunil Ahuja, M.D., South Texas Veterans Health Care System
  7400 Merton Minter Boulevard, San Antonio, TX 78229
- Kathrina Alexander, M.D., Veterans Health Care System of the Ozarks
  1100 North College Avenue, Fayetteville, AR 72703
- Xiao M. Androulakis, M.D., Columbia VA Health Care System
  6439 Garners Ferry Road, Columbia, SC 29209
- Prakash Balasubramanian, M.D., William S. Middleton Memorial Veterans Hospital
  2500 Overlook Terrace, Madison, WI 53705
- Zuhair Ballas, M.D., Iowa City VA Health Care System
  601 Highway 6 West, Iowa City, IA 52246-2208
- Elizabeth S. Bast, M.D., M.P.H., Miami VA Health Care System
  1201 NW 16th Street, 11 GRC, Miami FL 33125
- Jean Beckham, Ph.D., Durham VA Medical Center
  508 Fulton Street, Durham, NC 27705
- Sujata Bhushan, M.D., VA North Texas Health Care System
  4500 S. Lancaster Road, Dallas, TX 75216
- Edward Boyko, M.D., VA Puget Sound Health Care System
  1660 S. Columbian Way, Seattle, WA 98108-1597
- David Cohen, M.D., Portland VA Medical Center
  3710 SW U.S. Veterans Hospital Road, Portland, OR 97239
- Louis Dellitalia, M.D., Birmingham VA Medical Center
  700 S. 19th Street, Birmingham AL 35233
- Gerald Wayne Dryden, Jr., M.D., Ph.D., Louisville VA Medical Center
  800 Zorn Avenue, Louisville, KY 40206
- L. Christine Faulk, M.D., Robert J. Dole VA Medical Center
  5500 East Kellogg Drive, Wichita, KS 67218-1607
- Joseph Fayad, M.D., VA Southern Nevada Healthcare System
  6900 North Pecos Road, North Las Vegas, NV 89086
- Daryl Fujii, Ph.D., VA Pacific Islands Health Care System
  459 Patterson Rd, Honolulu, HI 96819
- Saib Gappy, M.D., John D. Dingell VA Medical Center
  4646 John R Street, Detroit, MI 48201
- Frank Gesek, Ph.D., White River Junction VA Medical Center
  163 Veterans Drive, White River Junction, VT 05009
- Michael Godschalk, M.D., Richmond VA Medical Center
  1201 Broad Rock Blvd., Richmond, VA 23249
- Jennifer Greco, M.D., Sioux Falls VA Health Care System
  2501 W 22nd Street, Sioux Falls, SD 57105
- Todd W. Gress, M.D., Ph.D., Hershel “Woody” Williams VA Medical Center
  1540 Spring Valley Drive, Huntington, WV 25704
- Samir Gupta, M.D., M.S.C.S., VA San Diego Healthcare System
  3350 La Jolla Village Drive, San Diego, CA 92161
- Salvador Gutierrez, M.D., Edward Hines, Jr. VA Medical Center
  5000 South 5th Avenue, Hines, IL 60141
- Mark Hamner, M.D., Ralph H. Johnson VA Medical Center
  109 Bee Street, Mental Health Research, Charleston, SC 29401
- John Harley, M.D., Ph.D., Cincinnati VA Medical Center
  3200 Vine Street, Cincinnati, OH 45220
- Daniel J. Hogan, M.D., Bay Pines VA Healthcare System
  10,000 Bay Pines Blvd Bay Pines, FL 33744
- Adriana Hung, M.D., M.P.H., VA Tennessee Valley Healthcare System
  1310 24th Avenue, South Nashville, TN 37212
- Robin Hurley, M.D., W.G. (Bill) Hefner VA Medical Center
  1601 Brenner Ave, Salisbury, NC 28144
- Pran Iruvanti, D.O., Ph.D., Hampton VA Medical Center
  100 Emancipation Drive, Hampton, VA 23667
- Frank Jacono, M.D., VA Northeast Ohio Healthcare System
  10701 East Boulevard, Cleveland, OH 44106
- Darshana Jhala, M.D., Philadelphia VA Medical Center
  3900 Woodland Avenue, Philadelphia, PA 19104
- Seema Joshi, M.D., F.A.C.P., ABOIM; VA Eastern Kansas Health Care System
  4101 S 4th Street Trafficway, Leavenworth, KS 66048
- Scott Kinlay, M.B.B.S., Ph.D., VA Boston Healthcare System
  150 S. Huntington Avenue, Boston, MA 02130
- Michael Landry, Ph.D., Southeast Louisiana Veterans Health Care System
  2400 Canal Street, New Orleans, LA 70119
- Peter Liang, M.D., M.P.H., VA New York Harbor Healthcare System
  423 East 23rd Street, New York, NY 10010
- Suthat Liangpunsakul, M.D., M.P.H., Richard Roudebush VA Medical Center
  1481 West 10th Street, Indianapolis, IN 46202
- Jack Lichy, M.D., Ph.D., Washington DC VA Medical Center
  50 Irving St, Washington, D. C. 20422
- Tze Shien Lo, M.D., Fargo VA Health Care System
  2101 N. Elm, Fargo, ND 58102
- C. Scott Mahan, M.D., Charles George VA Medical Center
  1100 Tunnel Road, Asheville, NC 28805
- Ronnie Marrache, M.D., VA Maine Healthcare System Center, Augusta, ME 04330
- Stephen Mastorides, M.D., James A. Haley Veterans’ Hospital
  13000 Bruce B. Downs Blvd, Tampa, FL 33612
- Kristin Mattocks, Ph.D., M.P.H., Central Western Massachusetts Healthcare System
  421 North Main Street, Leeds, MA 01053
- Paul Meyer, M.D., Ph.D., Southern Arizona VA Health Care System
  3601 S 6th Avenue, Tucson, AZ 85723
- Jonathan Moorman, M.D., Ph.D., James H. Quillen VA Medical Center
  Corner of Lamont & Veterans Way, Mountain Home, TN 37684
- Providencia Morales, R.N., Northern Arizona VA Health Care System
  500 Highway 89 North, Prescott, AZ 86313
- Timothy Morgan, M.D., VA Long Beach Healthcare System
  5901 East 7th Street Long Beach, CA 90822
- Maureen Murdoch, M.D., M.P.H., Minneapolis VA Health Care System
  One Veterans Drive, Minneapolis, MN 55417
- Eknath Naik, M.D., Ph.D., West Palm Beach VA Medical Center,
  7305 North Military Trail, West Palm Beach, FL 33410-6400
- James Norton, Ph.D., VA Health Care Upstate New York
  113 Holland Avenue, Albany, NY 12208
- Olaoluwa Okusaga, M.D., Michael E. DeBakey VA Medical Center
  2002 Holcombe Blvd, Houston, TX 77030
- Michael K. Ong, M.D., VA Greater Los Angeles Health Care System
  11301 Wilshire Blvd, Los Angeles, CA 90073
- Kris Ann Oursler, M.D., Salem VA Medical Center
  1970 Roanoke Blvd, Salem, VA 24153
- Ismene Petrakis, M.D., VA Connecticut Healthcare System
  950 Campbell Avenue, West Haven, CT 06516
- Samuel Poon, M.D., Manchester VA Medical Center
  718 Smyth Road, Manchester, NH 03104
- Amneet S. Rai, Pharm.D., VA Sierra Nevada Health Care System
  975 Kirman Avenue, Reno, NV 89502
- Michael Rauchman, M.D., St. Louis VA Health Care System
  915 North Grand Blvd, St. Louis, MO 63106
- Richard Servatius, Ph.D., Syracuse VA Medical Center
  800 Irving Avenue, Syracuse, NY 13210
- Satish Sharma, M.D., Providence VA Medical Center
  830 Chalkstone Avenue, Providence, RI 02908
- River Smith, Ph.D., Eastern Oklahoma VA Health Care System
  1011 Honor Heights Drive, Muskogee, OK 74401
- Peruvemba Sriram, M.D., N. FL/S. GA Veterans Health System
  1601 SW Archer Road, Gainesville, FL 32608
- Patrick Strollo, Jr., M.D., VA Pittsburgh Health Care System
  University Drive, Pittsburgh, PA 15240
- Neeraj Tandon, M.D., Overton Brooks VA Medical Center
  510 East Stoner Ave, Shreveport, LA 71101
- Philip Tsao, Ph.D., VA Palo Alto Health Care System
  3801 Miranda Avenue, Palo Alto, CA 94304-1290
- Gerardo Villareal, M.D., New Mexico VA Health Care System
  1501 San Pedro Drive, S.E. Albuquerque, NM 87108
- Jessica Walsh, M.D., VA Salt Lake City Health Care System
  500 Foothill Drive, Salt Lake City, UT 84148
- John Wells, Ph.D., Edith Nourse Rogers Memorial Veterans Hospital
  200 Springs Road, Bedford, MA 01730
- Jeffrey Whittle, M.D., M.P.H., Clement J. Zablocki VA Medical Center
  5000 West National Avenue, Milwaukee, WI 53295
- Mary Whooley, M.D., San Francisco VA Health Care System
  4150 Clement Street, San Francisco, CA 94121
- Peter Wilson, M.D., Atlanta VA Medical Center
  1670 Clairmont Road, Decatur, GA 30033
- Junzhe Xu, M.D., VA Western New York Healthcare System
  3495 Bailey Avenue, Buffalo, NY 14215-1199
- Shing Shing Yeh, Ph.D., M.D., Northport VA Medical Center
  79 Middleville Road, Northport, NY 11768
- Andrew W. Yen, M.D., VA Northern California Health Care System
  10535 Hospital Way, Mather, CA 95655

**Supplementary References**

1	Jaganathan, K. *et al.* Predicting Splicing from Primary Sequence with Deep Learning. *Cell* **176**, 535-548 e524 (2019). <https://doi.org:10.1016/j.cell.2018.12.015>

2	Dron, J. S. *et al.* Loss-of-Function *CREB3L3* Variants in Patients With Severe Hypertriglyceridemia. *Arteriosclerosis, Thrombosis, and Vascular Biology* **40**, 1935-1941 (2020). <https://doi.org:10.1161/atvbaha.120.314168>
